# Estimated durations of asymptomatic, symptomatic, and care-seeking phases of tuberculosis disease

**DOI:** 10.1101/2021.03.17.21253823

**Authors:** Chu-Chang Ku, Peter MacPherson, McEwen Khundi, Rebecca Nzawa, Helena RA. Feasey, Marriott Nliwasa, Katherine C. Horton, Elizabeth L. Corbett, Peter J. Dodd

**Affiliations:** School of Health and Related Research, University of Sheffield, United Kingdom; Malawi-Liverpool-Wellcome Trust Clinical Research Programme, Queen Elizabeth Central Hospital, Blantyre, Malawi; Department of Clinical Sciences,Liverpool School of Tropical Medicine, Liverpool, UK; Department of Clinical Research, London School of Hygiene and Tropical Medicine, London, UK; Helse Nord TB Initiative, University of Malawi College of Medicine, Malawi; Department of Infectious Disease Epidemiology, London School of Hygiene and Tropical Medicine, London, UK

## Abstract

Ratios of bacteriologically-positive tuberculosis prevalence to notification rates are used to characterise typical durations of tuberculosis disease, but have not accounted for asymptomatic periods prior to care-seeking. We developed novel statistical models to estimate progression from initial bacteriological-positivity including smear conversion, symptom onset and initial care-seeking and fitted them to tuberculosis prevalence survey and notification data (one subnational and 11 national datasets) within a Bayesian framework. Asymptomatic tuberculosis duration was in the range 4 – 8 months for African countries; three countries in Asia showed longer durations of > 1 year. Care-seeking typically began half-way between symptom onset and notification. Our method also estimated smear progression rates and case-detection ratios. We found evidence for higher case-detection ratios and much shorter durations of tuberculosis for people living with HIV. To eradicate tuberculosis transmission, greater gains may be achieved by proactively screening people without symptoms through active case finding interventions.

## Introduction

Population surveys of the prevalence of bacteriologically-positive tuberculosis (TB) disease are a key tool for understanding TB epidemiology and burden, and, when repeated over time, for monitoring the impacts of control efforts. Over the last decade, the World Health Organization (WHO) has encouraged and facilitated a series of nationally-representative TB prevalence surveys in priority countries [1]. TB prevalence surveys are a particularly important source of data for estimating TB incidence in high-burden settings where notification systems are imperfect [1], and while typically powered to achieve a 20% relative precision in the measurement of TB prevalence [2], surveys also contain additional information on subgroups which has, for instance, highlighted the higher burden of TB among men [3].

By comparing prevalence with notifications—usually as a prevalence-to-notification (P:N) ratio—one can estimate a typical timescale for prevalent TB, the inverse of which (the patient diagnostic rate) provides a measurable indicator of the effectiveness of case detection [4]. Comparing P:N ratios between sexes has shown men have poorer access to care in many settings [3]. However, the influence of age and HIV have not been analysed.

Several prevalence surveys also record whether individuals with TB were symptomatic and some record whether individuals with TB had previously sought care for their symptoms [5]. These surveys have found that large proportions of cases—around half—do not report symptoms [6,7]. Lack of symptoms among those with microbiologically-confirmed pulmonary TB has contributed to an increased understanding of asymptomatic and subclinical TB as being part of a spectrum of TB disease [8], and a potentially important contributor to TB burden and transmission [7,9].

Many aspects of the natural history of TB disease prior to (or without) treatment remain very uncertain because ethical considerations mean we must rely on historic data from the pre-chemotherapy era. For example, how often and how quickly individuals with smear-negative TB progress to smear-positive disease is unclear. Similarly, while there are data to suggest a typical duration of around three years for untreated TB disease [10], there is only weak and indirect data to quantify the duration of TB among people living with HIV (PLHIV), which is thought to be much shorter [11].

A better understanding of the duration of asymptomatic bacteriologically-positive pulmonary TB could inform how screening, case finding, and prevention interventions are designed and implemented. We therefore sought to leverage prevalence survey data from a variety of settings to investigate the duration of asymptomatic TB disease, and typical delays to care-seeking and notification. We used a novel Bayesian framework within which we incorporated uncertainty, disease progression before detection, and trends in incidence. This approach also provided estimates of TB incidence, incidence trends, case-detection ratios, and for some settings, stratification by age and HIV infection status.

## Methods

### Study populations and data

#### TB prevalence surveys

We analysed data from eleven national and one sub-national settings with a TB prevalence survey conducted after 2010. We excluded TB prevalence surveys that relied solely on symptom screening to decide who to sample for bacteriological testing and restricted to the population aged 15 years or older.

The eleven national settings with recent TB prevalence surveys are Cambodia [12], Ethiopia [13], Kenya [14], Lao PDR [15], Malawi [16], Pakistan [17], Philippines [18], Tanzania [5,19], Uganda[20], Vietnam [21], and Zambia [22]. For those prevalence surveys, we extracted the raw counts of eligible subjects and active cases, excluding those already on TB treatment if possible. Seven surveys included healthcare behaviour data enabling us to identify whether symptomatic cases had started seeking care.

Summaries of prevalence survey data are shown in **Table 1**. The proportion of prevalent cases that was asymptomatic ranged from 30% in Malawi, to 70% in Cambodia, and the proportion of prevalent cases that were sputum smear positive varied from 23% in Vietnam to 84% in Tanzania. Definitions used for symptoms, criteria used for bacteriological testing, and bacteriological testing methods are summarized in **Table 1-table supplement 1**.

**Table 1.** Epidemiological indices and data

|  | Cambodia | Ethiopia | Kenya | Lao PDR | Malawi | Pakistan | Philippines | Uganda | Tanzania | Viet Nam | Zambia |
| --- | --- | --- | --- | --- | --- | --- | --- | --- | --- | --- | --- |
| Population aged 15 and above, 2019, million | 11.4 | 66.9 | 32 | 4.9 | 10.5 | 140.6 | 75.2 | 23.7 | 32.6 | 74.1 | 9.9 |
| **Prevalence survey** |  |  |  |  |  |  |  |  |  |  |  |
| Survey year | 2011 | 2010-2011 | 2016 | 2010-2011 | 2013 | 2010-2011 | 2016 | 2014-2015 | 2012 | 2017-2018 | 2013-2014 |
| Prevalence in the survey year, per 100,000* | 839 (748-933) | 236 (193-281) | 478 (425-532) | 604 (528-683) | 418 (348-491) | 325 (291-360) | 998 (908-1090) | 395 (335-456) | 305 (258-355) | 358 (311-406) | 575 (508-644) |
| **WHO data/estimates** |  |  |  |  |  |  |  |  |  |  |  |
| Case notification, 2019, per 100,000 | 247 (244-250) | 150 (149-150) | 238 (236-240) | 138 (135-142) | 146 (144-148) | 201 (200-202) | 488 (486-489) | 238 (236-240) | 212 (210-213) | 136 (135-137) | 340 (336-343) |
| Case-detection ratio | 57 (40.0 – 88) | 69 (51.0 – 98) | 63 (43.0 – 100) | 57 (40.0 – 89) | 56 (35.0 – 110) | 64 (48.0 – 90) | 63 (40.0 – 110) | 65 (43.0 – 110) | 53 (30.0 – 110) | 57 (40.0 – 90) | 58 (41.0 – 90) |
| Incidence rate, 2009, per 100,000 | 387 (220 – 546) | 208 (133 – 283) | 385 (194 – 572) | 206 (120 – 288) | 218 (85 – 361) | 351 (225 – 478) | 705 (310 – 1102) | 304 (135 – 473) | 356 (98 – 617) | 219 (127 – 312) | 534 (302 – 766) |
| **Model estimates: All TB regardless of symptomatic or not** |  |  |  |  |  |  |  |  |  |  |  |
| Duration by P:N ratio, month** | 28.9 (25.8 – 32.3) | 10.2 (8.4 – 12.3) | 25.1 (22.5 – 28.0) | 109.4 (95.1 – 125.6) | 29.7 (25.1 – 34.9) | 17.8 (15.9 – 19.9) | 33.3 (30.5 – 36.4) | 24.5 (21.0 – 28.5) | 20.8 (17.7 – 24.2) | 33.0 (28.6 – 37.3) | 16.0 (14.2 – 18.0) |
| Time to notification (Total duration), month | 22.0 (18.9 – 25.8) | 9.0 (7.5 – 10.8) | 18.3 (16.1 – 21.0) | 36.1 (29.9 – 45.3) | 21.8 (18.4 – 25.9) | 13.1 (11.5 – 14.8) | 22.3 (19.6 – 25.6) | 19.1 (16.4 – 22.2) | 16.2 (14.0 – 18.8) | 20.7 (17.8 – 24.5) | 13.4 (11.8 – 15.2) |
| Case-detection ratio*** | 67% (56% – 79%) | 87% (80% – 94%) | 69% (60% – 79%) | 24% (19% – 31%) | 63% (53% – 74%) | 76% (68% – 84%) | 53% (46% – 62%) | 63% (53% – 72%) | 68% (59% – 77%) | 59% (50% – 70%) | 79% (71% – 88%) |
| Incidence rate, per 100,000 | 342 (285 – 403) | 168 (156 – 183) | 360 (312 – 414) | 595 (449 – 743) | 222 (187 – 265) | 287 (259 – 319) | 914 (782 – 1,055) | 354 (305 – 412) | 307 (271 – 352) | 230 (194 – 272) | 426 (382 – 474) |
| **Model estimates: All symptomatic TB** |  |  |  |  |  |  |  |  |  |  |  |
| Duration by P:N ratio, month | 8.7 (7.1 – 10.5) | 5.3 (4.0 – 6.7) | 11.8 ( 9.9 – 13.8) | 53.4 (43.6 – 64.1) | 20.6 (16.7 – 24.9) | 10.9 (9.4 – 12.4) | 10.9 (9.2 – 12.7) | 11.9 (9.4 – 14.6) | 13.3 (10.7 – 16.0) | 21.9 (18.6 – 25.6) | 9.7 (8.2 – 11.3) |
| Time to care-seeking start, month | 1.2 (0.6 – 1.9) | N.A. | 5.6 (4.6 – 6.8) | N.A. | 6.6 (4.8 – 8.6) | N.A. | 4.2 (3.3 – 5.2) | 4.2 (3.0 – 5.6) | 6.3 (4.9 – 7.8) | N.A. | 4.1 (3.3 – 5.1) |
| Time to notification, month | 8.0 (6.7 – 9.6) | 4.8 (3.7 – 6.0) | 9.5 (8.2 – 11.1) | 21.6 (18.0 – 26.8) | 15.8 (13.2 – 19.0) | 8.2 (7.2 – 9.4) | 9.2 (7.9 – 10.6) | 10.4 (8.5 – 12.5) | 10.9 (9.1 – 13.0) | 14.2 (12.1 – 16.8) | 8.5 (7.3 – 9.8) |
| Case-detection ratio | 83% (77% – 89%) | 92% (87% – 96%) | 80% (73% – 86%) | 36% (29% – 44%) | 69% (59% – 79%) | 82% (76% – 88%) | 75% (69% – 81%) | 75% (67% – 82%) | 75% (67% – 82%) | 66% (57% – 75%) | 85% (78% – 91%) |
| Incidence rate, per 100,000 | 270 (252 – 292) | 160 (154 – 169) | 309 (286 – 338) | 396 (315 – 482) | 203 (177 – 235) | 265 (247 – 285) | 643 (593 – 700) | 296 (269 – 329) | 277 (252 – 309) | 206 (181 – 235) | 397 (369 – 429) |
| **Other estimates / metrics** |  |  |  |  |  |  |  |  |  |  |  |
| Annual decline rate , % per year | 4.7% (4.4% – 5.0%) | 7.3% (7.1% – 7.5%) | 1.5% (1.3% – 1.6%) | -8.0% (-8.5% – -7.5%) | 2.9% (2.6% – 3.2%) | 1.6% (1.5% – 1.7%) | -8.3% (-8.4% – -8.2%) | -2.4% (-2.7% – -2.2%) | -1.3% (-1.5% – -1.1%) | 1.0% (0.7% – 1.4%) | 4.9% (4.7% – 5.1%) |
| Asymptomatic phase over total duration, % | 64% (58% – 69%) | 47% (39% – 56%) | 48% (42% – 53%) | 40% (35% – 45%) | 27% (21% – 35%) | 37% (32% – 42%) | 59% (53% – 65%) | 45% (38% – 53%) | 32% (26% – 40%) | 31% (26% – 37%) | 37% (31% – 43%) |
| Symptomatic TB without care-seeking, % | 15% (9% – 23%) | N.A. | 59% (51% – 67%) | N.A. | 42% (33% – 51%) | N.A. | 46% (38% – 54%) | 41% (31% – 50%) | 57% (48% – 67%) | N.A. | 48% (40% – 56%) |
| Self-cure before case-detection | 32% (20% – 42%) | 14% (8% – 21%) | 27% (16% – 36%) | 50% (35% – 61%) | 30% (19% – 41%) | 20% (12% – 28%) | 32% (20% – 42%) | 27% (17% – 37%) | 23% (14% – 33%) | 30% (19% – 41%) | 20% (12% – 28%) |
| Deaths before case-detection | 7% (6% – 9%) | 3% (2% – 4%) | 6% (5% – 7%) | 19% (15% – 26%) | 10% (8% – 12%) | 6% (5% – 7%) | 6% (5% – 7%) | 8% (6% – 10%) | 8% (6% – 9%) | 12% (10% – 14%) | 5% (4% – 6%) |

\* values are crude rates or proportions with 95% Binomial confidence intervals, while the model estimates are mean estimators with 95% credible intervals
\*\* P:N ratios use prevalence from the model estimates for 2019 and notified cases from WHO 2019 TB notification data
\*\*\* Case-detection ratio is defined as the ratio of notified cases and the estimated incident cases in percentage

The sub-national TB prevalence survey from Blantyre, Malawi and the national survey from Kenya [15] included individual data, allowing analysis by smear status, age, and HIV status [14]. In Blantyre, Malawi, the prevalence survey was conducted as part of a community cluster randomised trial across approximately 750,000 adults in 72 densely populated neighbourhoods of the city. For Blantyre, we aligned symptom definition with that of Kenya by considering cough of >=2 weeks, considering cough of any duration in a sensitivity analysis.

#### Definitions

Unless otherwise stated, we used the definition of TB symptoms and health-care seeking adopted by each TB prevalence survey. Asymptomatic TB was taken to be bacteriologically-positive pulmonary TB in those reporting no TB symptoms, and we assumed no health-care seeking for TB during this phase. Participants with prevalent TB who were already taking TB treatment were excluded. We assumed TB treatment initiation, TB confirmation, and case notification are identical events for the modelling.

#### TB notification data

We used TB notification data and treatment outcome data from the WHO TB database for the countries with national prevalence surveys [12–22]. We included new and relapse TB regardless of smear status since 2013. We applied the proportions of HIV among TB notifications by sex, age group, and smear status to disaggregate the WHO notification data. Where notification data exhibited level shifts suggestive of changes in reporting, only consistent, sequential data were used.

For Kenya, we obtained data simultaneously stratified by HIV and age group from the National Tuberculosis Programme. For Blantyre, we obtained individual-level notification data from an enhanced surveillance system. We matched the age-groups of the Kenya data to the finest scale of WHO case notification data; for Blantyre, Malawi, with lower case counts, we used two age groups (15 to 49, and 50+ years).

#### Demographic data

In order to model mortality and project national numbers of prevalent cases to relate to notifications, we used age- and sex-specific background mortality rates and population sizes from 2019 World Population Prospect (WPP) data, using the mid-year population estimates [23]. For Blantyre, we rescaled the Malawi demographic data to local 2008 and 2018 population censuses [24,25].

The demographic and notification data to replicate the analysis are available at **Source Data**.

### Statistical methods

#### Estimation of TB progression and care-seeking behaviour

We developed three state transition models of TB case-detection to match available data, and fitted them to estimate parameters driving TB progression and care-seeking behaviour. **Figure 1** shows that all models contain an asymptomatic phase and progress to the symptomatic phase. Model A presents a basic transition between asymptomatic/symptomatic phases; Model B divides the symptomatic phase by presence of care-seeking intentions, and Model C details the conversion between smear-negative and smear-positive and care-seeking by smear status. Model B is only applicable to the prevalence surveys that reported care-seeking behaviours. We did not combine Model B and C because the symptoms, smear status, and care-seeking behaviours prevalence surveys were not cross-tabulated in the published reports. We constructed a likelihood depending on these states to fit to TB prevalence and case-notification data. For Kenya and Blantyre data with Model C, we incorporated proportional hazard models relating the rate of symptom onset and the care-seeking rates by smear status to covariates of age, sex, and HIV status.

**Figure 1:**
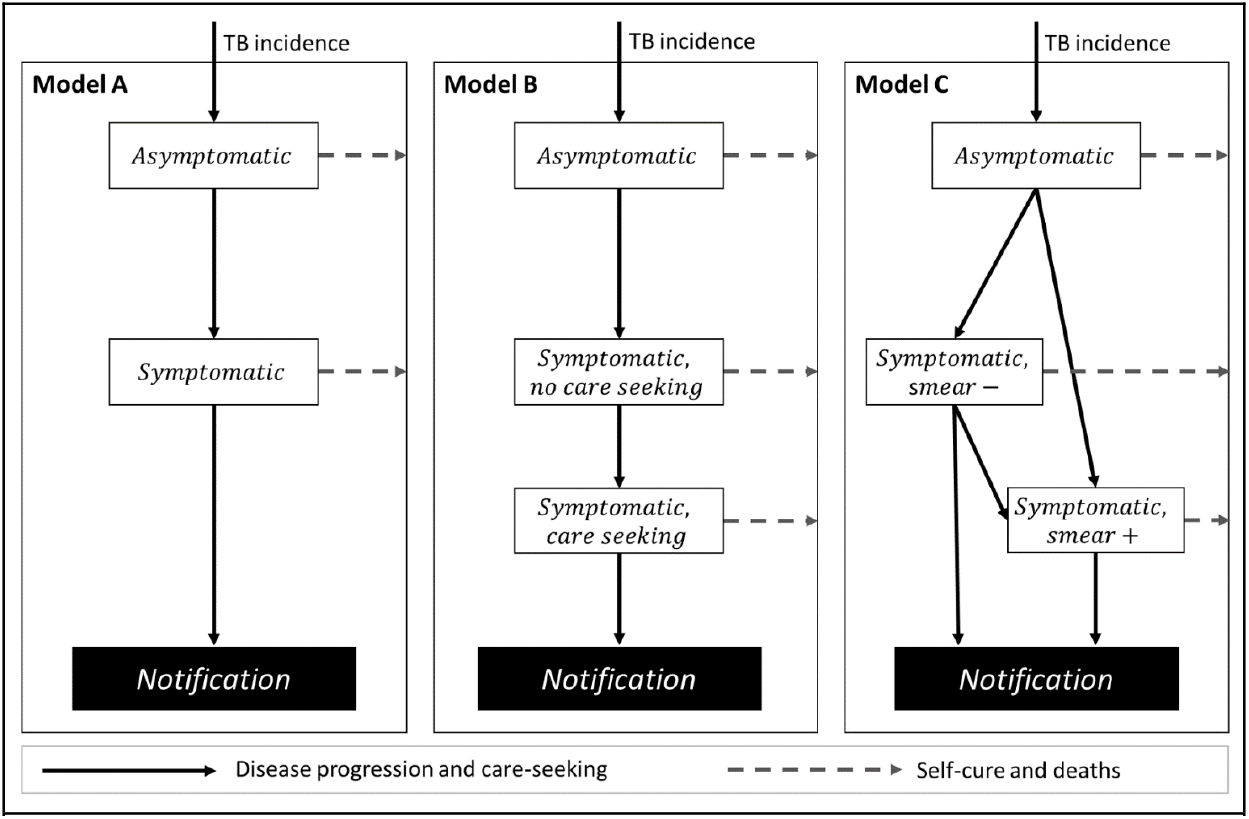
Diagrams of model structure. White boxes are all bacteriologically-positive TB accessible to prevalence surveys, with structure matched to available data. Models are additionally stratified by sex, and for Kenya and Blantyre, Malawi by age and HIV in addition.

We calculated a single weighted-mean background mortality rate from WPP data in each country, using WHO TB notifications as weights. For the analyses with HIV stratification, we added HIV-related deaths for PLHIV from the UNAIDS database: 0.016 per year (21,000 / 1,300,000) for Kenya in 2016 and 0.011 per year (11,000 / 1,000,000) for Blantyre (based on Malawi data). In asymptomatic TB, we assumed no TB-related deaths, but did allow self-cure and deaths due to other causes. Smear status-specific TB death rates were applied to symptomatic TB states based on Tiemersma et al. [10]. For Model A and Model B, regardless of smear status, we applied the untreated TB deaths weighted by smear status in the notification data. We conducted sensitivity analyses for assumed excess mortality rates of asymptomatic/symptomatic TB. We also considered two extreme assumptions for HIV-TB excess mortality: the same TB mortality rate as HIV-negative people, and an excess mortality rate of 5% per month as considered in Vynnycky et al. [26] for gold miners in South Africa.

We assumed each state steadily declined with a constant shared rate. The statistical models were constructed within a Bayesian framework; **Figure 1-figure supplement 1-3** details the mathematical formulation and priors. We fitted all models by Markov chain Monte Carlo (MCMC) using R with rstan. MCMC runs all converged with Gelman-Rubin R^2 statistics < 1.05. Inferences were based on 3,000 samples from three chains after burn-in. Processed data and all source code for this analysis are available on Github as [https://github.com/TimeWz667/AsymTB.git].

#### Metrics calculated

For each setting, we used posterior samples to calculate: the TB incidence; the mean times from incident disease to development of symptoms (i.e. the duration of asymptomatic disease), to case-seeking initiation (where applicable), and to notification; the case detection ratio (CDR) as the ratio of incidence and notification rates; and the proportion of TB cases reaching each stage of care. Aggregated quantities were computed weighted by model-estimated incidence. We also output a joint posterior for the proportion of symptomatic cases initially smear positive and the subsequent smear-conversion rate. We report means and 95% credible intervals (CrIs).

### Ethical approval

The Blantyre, Malawi TB prevalence survey received ethical approval from the College of Medicine, University of Malawi, and from London School of Hygiene and Tropical Medicine.

## Results

### Duration of asymptomatic disease, time to care-seeking initiation and time to case-detection

The national estimates for duration of asymptomatic disease typically ranged around 3-8 months. However, three countries in Asia (Lao PDR, Cambodia, Philippines) showed longer durations of over one year (see **Figure 2**). These countries also had long total durations: 22 months for Cambodia and Philippines, and three years for Lao PDR. Only one country (Ethiopia) had a total duration lower than 12 months. In the seven countries where we could estimate the delay to care-seeking initiation, apart from the Philippines (16 months), delay varied between 1.2 (0.7 – 1.9) months for Cambodia and 6.6 (95% CrI: 4.8 – 8.6) months for Malawi. The asymptomatic phase represented between 27% and 63% of time as a prevalent case, and treatment initiation occurred between 15% and 59% of the way between symptom onset and notification (or death or self-cure).

**Figure 2.**
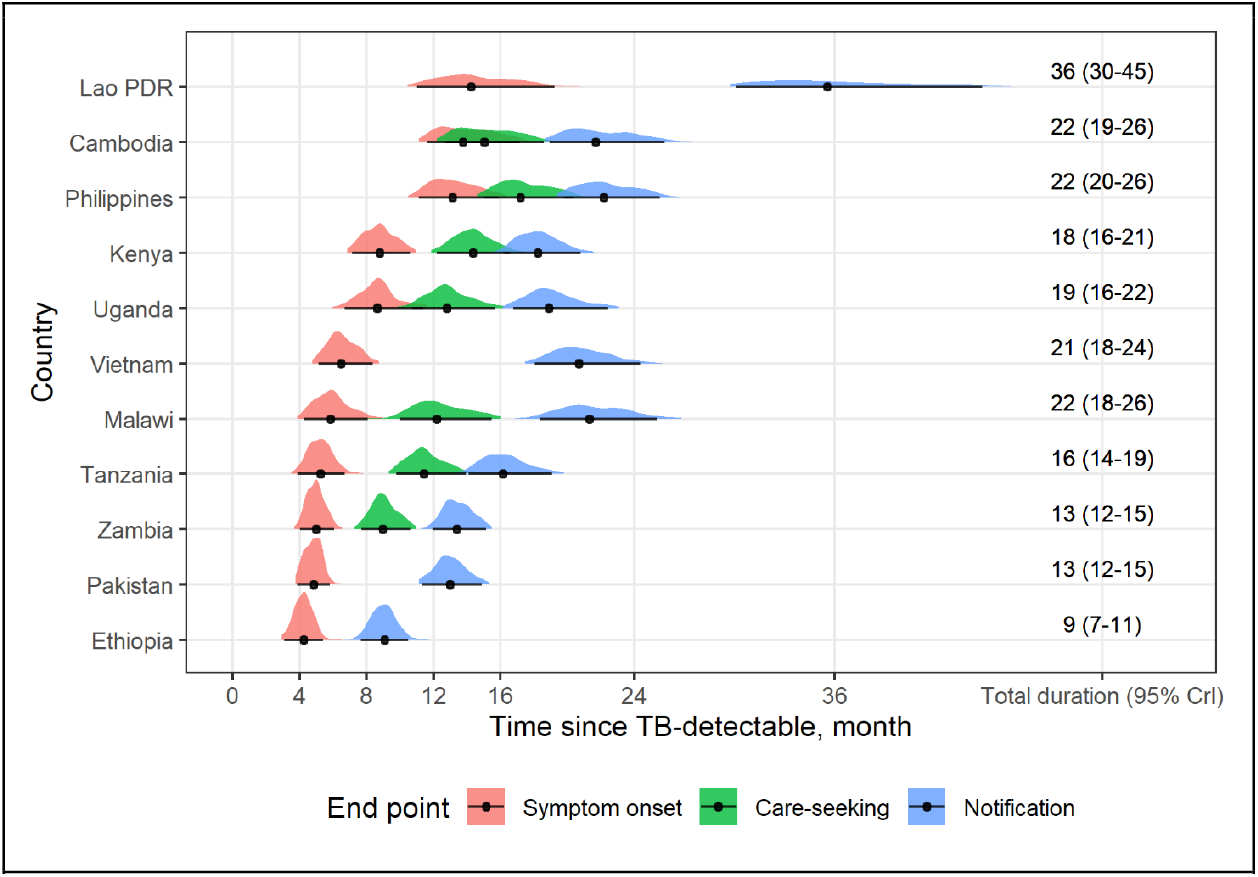
Time to events during bacteriologically-positive TB disease. Times are measured from bacteriologically-positive TB disease incidence in months. Median and 95% quantiles are shown as points and error bars, respectively. Posterior distributions are shown by colored kernel density estimates.

### Development of smear-positivity

**Figure 3** shows the joint posterior probability densities of smear conversion rates and the proportion of symptomatic cases that were smear-positive at symptom onset. Estimates of smear conversion rates were linearly related to the initial proportion of symptomatic cases that were smear-positive. Pooling weighted by TB notifications in 2019 (excluding Malawi and Vietnam as outliers) yielded a gradient of 0.239 years, and a pooled intercept of 53.4% smear positive (for zero smear conversion rate).

**Figure 3.**
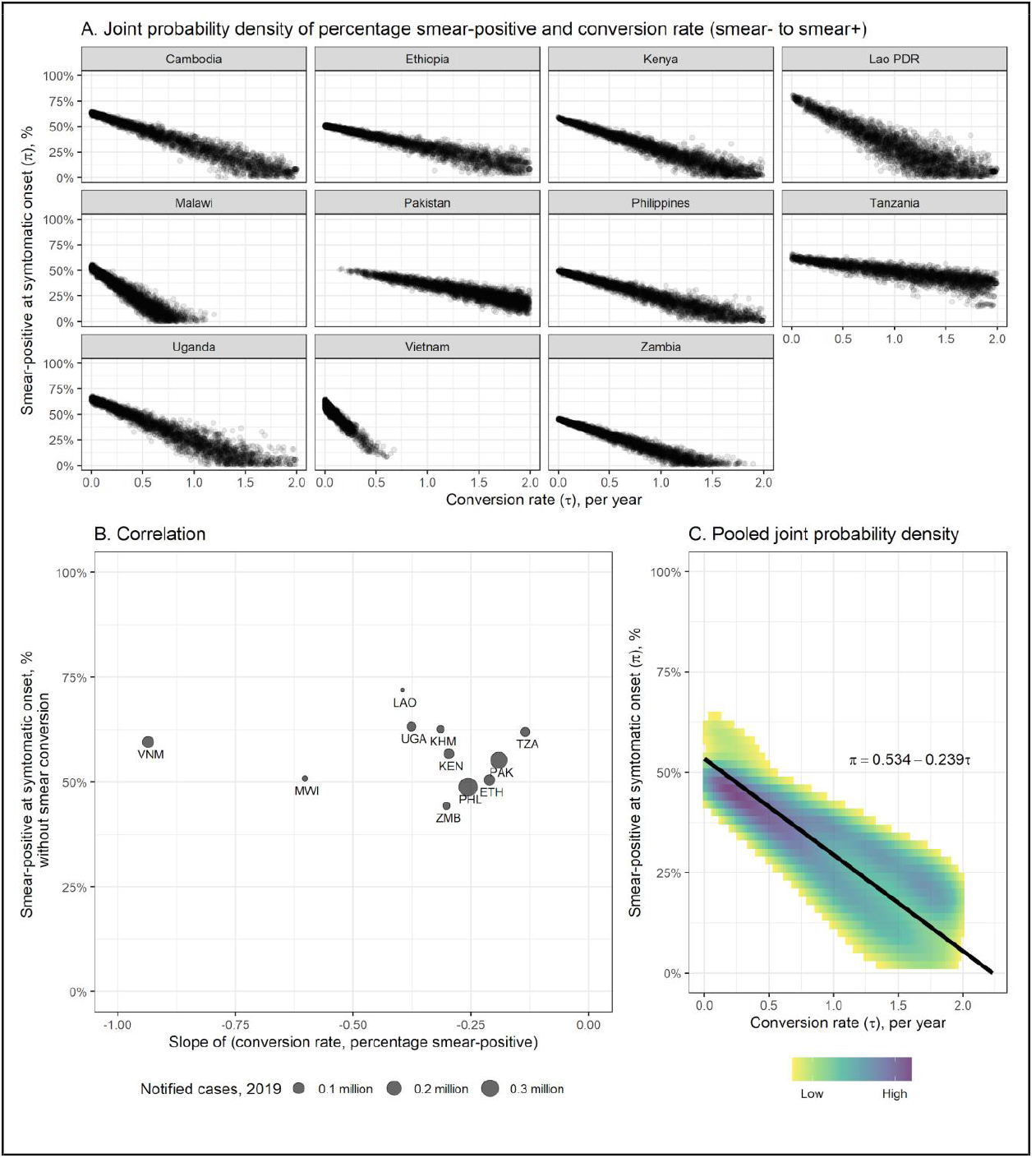
Smear-conversion rate and initial proportion smear-positive in symptomatic bacteriologically-positive TB disease. (A) joint posterior probability densities of initial proportion smear-positive (y-axis) and hazard of converting from smear-negative to smear-positive (x-axis) in symptomatic bacteriologically-positive TB by country, based on Model 3. (B) the correlation of percentage smear-positive at symptom onset and smear-type conversion rate. X-axis indicates the slopes of Fig 3-A estimated by linear regression (KHM=Cambodia, ETH=Ethiopia, KEN=Kenya, LAO=Lao People’s Democratic Republic, MWI=Malawi, PAK=Pakistan, PHL= Philippines, TZA=United Republic of Tanzania, UGA=Uganda, VNM=Viet Nam, ZMB=Zambia) (C) joint probability density of initial proportion smear-positive (y-axis) and hazard of converting from smear-negative to smear-positive (x-axis) pooled by weights of notified cases in 2019 (excluding Malawi and Vietnam as outliers).

### Estimates of epidemiological indices considering asymptomatic phase of TB

**Table 1** shows the empirical estimates of total duration and total asymptomatic duration based on P:N ratios. Empirical estimates of duration systematically overestimate the durations, because they implicitly assume all episodes of TB disease end in notification. Modelling self-cure and death leads to shorter estimates of duration and differences in estimated CDRs from WHO estimates. The proportions of incident TB who die or self-cure before being detected are shown in **Table 1**. Estimates of TB incidence are also distinct but comparable to WHO incidence estimates. We estimated rates of TB incidence change from a decline of 7.3% (95% CrI: 7.1% – 7.5%) per year in Ethiopia up to an increase of 8.3% (95% CrI: 8.2% – 8.4%) per year in the Philippines.

### Care-seeking cascade estimates

**Figure 4** shows care cascade estimates for the year of national prevalence surveys, showing the proportion of incident TB cases that develop symptoms, begin seeking care, initiate treatment, and finally successfully complete treatment. There were notable differences between countries, with the proportion initiating TB treatment ranging from 30% (95% CrI: 24% – 38%) in Lao PDR to 83% (95% CrI: 72% – 88%) in Ethiopia. Versions of these figures with cohort timings are shown in **Figure 4-figure supplement 1**.

**Figure 4.**
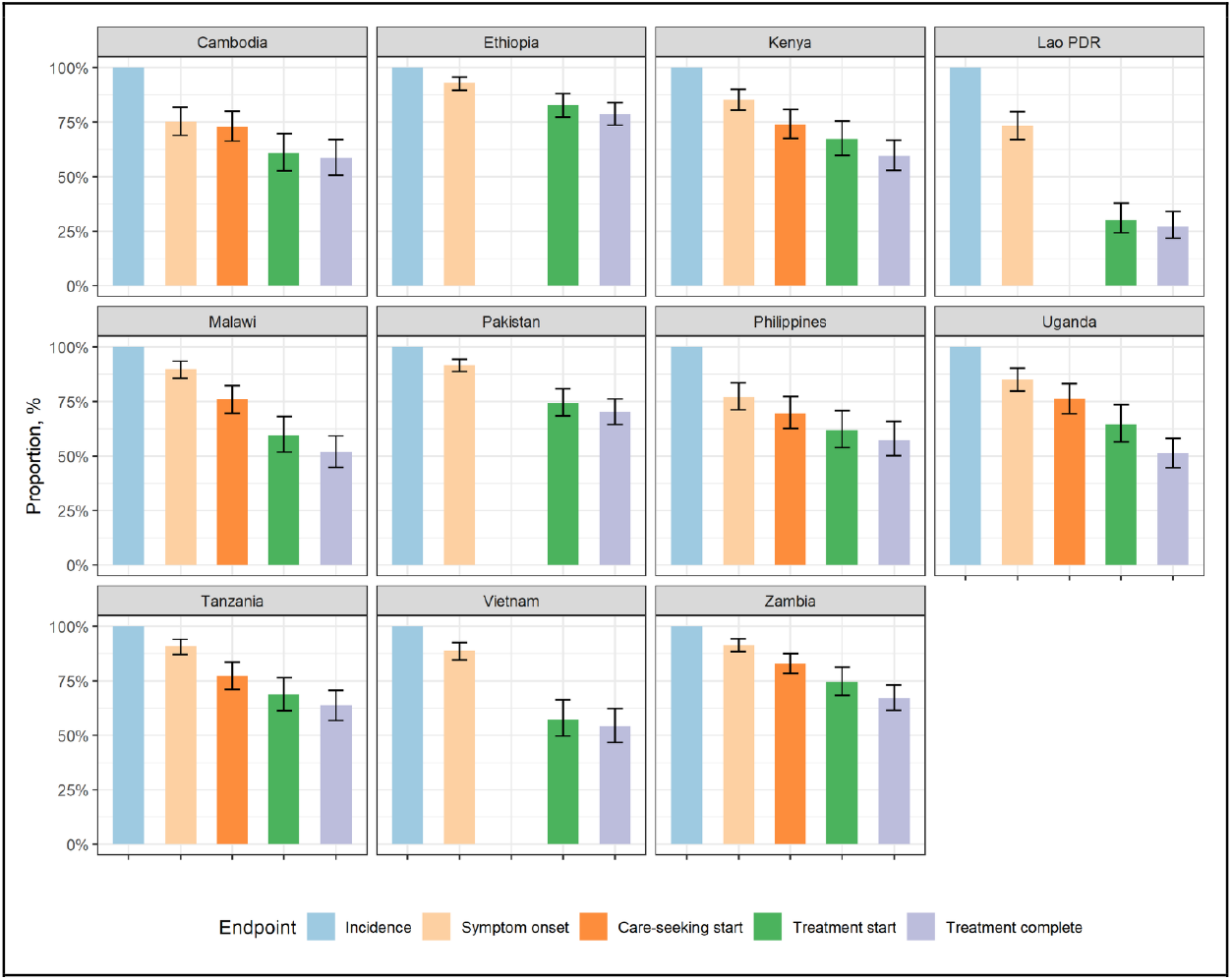
Healthcare cascade. The values are the fraction of the incident cohort reaching each stage. The second bars may be missing if the TB prevalence survey did not report results on care-seeking behaviour.

### Sex, age, and HIV

For Kenya and Blantyre, Malawi, we were able to examine results stratified by sex, age, and HIV-status (see **Table 2**). For Kenya we found total durations of 18.8 months (95% CrI: 16.1 – 21.8) for men and 15.4 months (95% CrI: 13.0 – 18.1) for women, and for Blantyre 9.9 months (95% CrI: 6.7 – 13.8) for men and 6.6 months (95% CrI: 3.5 – 10.2) for women, in line with previous work suggesting longer durations for men [3].

**Table 2.** Asymptomatic, symptomatic and total TB duration by sex, age and HIV status

|  |  |  | Asymptomatic<br>duration, month | Symptom onset rate after TB activation, per year |  |  | Symptomatic<br>duration, month | Total<br>duration, month |
| --- | --- | --- | --- | --- | --- | --- | --- | --- |
|  |  |  |  | Crude rate | Crude RR | Adjusted RR |  |  |
| Kenya | Overall |  | 8.3 (6.8 – 10.1) | 1.25 (1.05 – 1.50) |  |  | 8.4 (7.1 – 9.7) | 16.7 (14.5 – 19.1) |
|  | Sex | Female | 8.1 (6.2 – 10.3) | 1.30 (1.01 – 1.69) | reference | reference | 7.2 (5.8 – 8.8) | 15.4 (13.0 – 18.1) |
|  |  | Male | 9.0 (7.2 – 11.2) | 1.14 (0.93 – 1.40) | 0.89 (0.63 – 1.20) | 0.68 (0.50-0.89) | 9.7 (8.2 – 11.5) | 18.8 (16.1 – 21.8) |
|  | Age | [15,25) | 9.5 (6.6 – 13.0) | 1.11 (0.76 – 1.60) | reference | reference | 10.4 (7.9 – 13.2) | 20.0 (16.1 – 24.4) |
|  |  | [25,35) | 8.6 (6.3 – 11.4) | 1.23 (0.90 – 1.66) | 1.13 (0.75 – 1.67) | 1.20 (0.77-1.69) | 9.4 (7.5 – 11.5) | 18.0 (15.0 – 21.6) |
|  |  | [35,45) | 7.1 (5.0 – 9.7) | 1.55 (1.08 – 2.22) | 1.43 (0.87 – 2.17) | 1.72 (1.10-2.54) | 7.7 (5.9 – 9.7) | 14.7 (11.9 – 18.1) |
|  |  | [45,55) | 7.4 (4.8 – 10.7) | 1.49 (0.94 – 2.31) | 1.39 (0.80 – 2.43) | 1.82 (0.95-3.17) | 8.3 (6.0 – 11.1) | 15.7 (12.4 – 19.7) |
|  |  | [55,65) | 9.6 (6.1 – 13.7) | 1.10 (0.70 – 1.74) | 1.02 (0.55 – 1.72) | 1.23 (0.68-2.06) | 5.2 (3.1 – 8.3) | 14.8 (10.9 – 19.4) |
|  |  | [65,Inf) | 7.1 (4.6 – 10.2) | 1.52 (0.97 – 2.35) | 1.43 (0.80 – 2.55) | 1.42 (0.89-2.34) | 5.3 (3.4 – 7.7) | 12.4 (9.4 – 16.0) |
|  | HIV | Non-HIV | 9.8 (7.9 – 12.2) | 1.03 (0.85 – 1.25) | reference | reference | 9.9 (8.5 – 11.7) | 19.7 (17.1 – 23.0) |
|  |  | PLHIV | 3.8 (2.4 – 5.6) | 3.09 (1.96 – 4.80) | 2.88 (1.82 – 4.54) | 2.45 (1.45-3.68) | 4.0 (2.5 – 5.7) | 7.8 (5.8 – 10.2) |
| Blantyre | Overall |  | 3.8 (2.3 – 5.8) | 3.10 (1.86 – 4.98) |  |  | 3.7 (2.3 – 5.4) | 7.5 (5.4 – 10.1) |
|  | Sex | Female | 3.5 (1.3 – 6.2) | 3.90 (1.73 – 8.83) | reference | reference | 3.1 (1.3 – 5.5) | 6.6 (3.5 – 10.2) |
|  |  | Male | 4.9 (2.6 – 7.9) | 2.46 (1.34 – 4.48) | 0.82 (0.29 – 1.82) | 0.81 (0.30-1.71) | 5.0 (2.9 – 7.7) | 9.9 (6.7 – 13.8) |
|  | Age | [15,50) | 4.0 (2.3 – 6.2) | 3.00 (1.74 – 5.03) | reference | reference | 3.7 (2.1 – 5.6) | 7.7 (5.3 – 10.3) |
|  |  | [50,Inf) | 3.5 (1.1 – 7.1) | 4.17 (1.46 – 10.84) | 1.53 (0.45 – 3.94) | 1.41 (0.47-4.16) | 4.6 (1.9 – 8.1) | 8.1 (4.3 – 12.5) |
|  | HIV | Non-HIV | 8.5 (5.0 – 12.7) | 1.29 (0.76 – 2.16) | reference | reference | 7.1 (4.3 – 10.8) | 15.6 (11.3 – 20.8) |
|  |  | PLHIV | 1.3 (0.3 – 3.0) | 12.79 (3.77 – 41.16) | 5.33 (1.90 – 11.92) | 4.81 (1.65-12.82) | 2.4 (1.0 – 4.1) | 3.7 (1.9 – 6.0) |

RR: risk ratio, PLHIV: people living with HIV

Adjusted results for Kenya suggested a longer asymptomatic phase in men than women; however, there were no clear differences in unadjusted results and results from Blantyre (see risk ratios in Table 2). There were also no clear patterns with respect to age. However, HIV status had a strong effect, with asymptomatic and symptomatic durations being shorter in PLHIV than those that were HIV negative. The total duration in PLHIV was 3.7 months (95% CrI: 1.9 – 6.0) for Blantyre and 7.8 months (95% CrI: 5.8 – 10.2) for Kenya.

We found that CDRs were consistently higher in PLHIV than in those without HIV infection, even with higher deaths rates for PLHIV with TB assumed (**Table 3**).

**Table 3.**
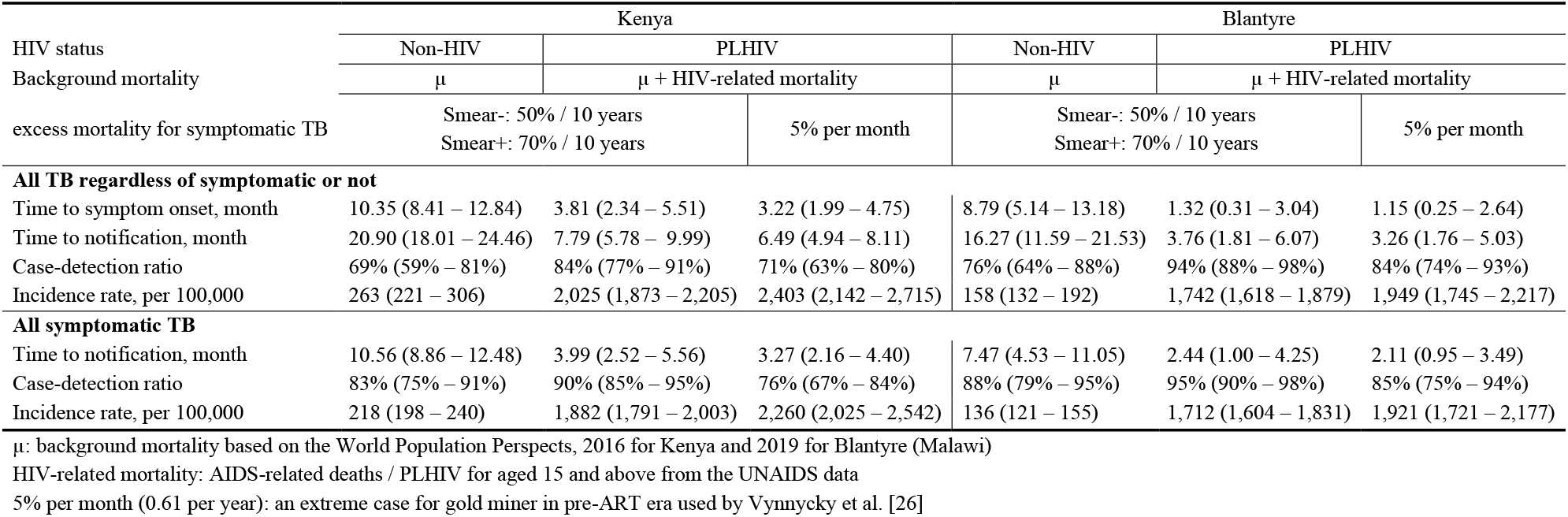
TB incidence, case detection ratio, and symptomatic duration by HIV status: Kenya and Blantyre, Malawi

Without HIV-specific case notification data at the national scale, **Figure 5** demonstrates the relationship between the asymptomatic duration estimates and the PLHIV among case-notification. We found no clear pattern of the association.

**Figure 5.**
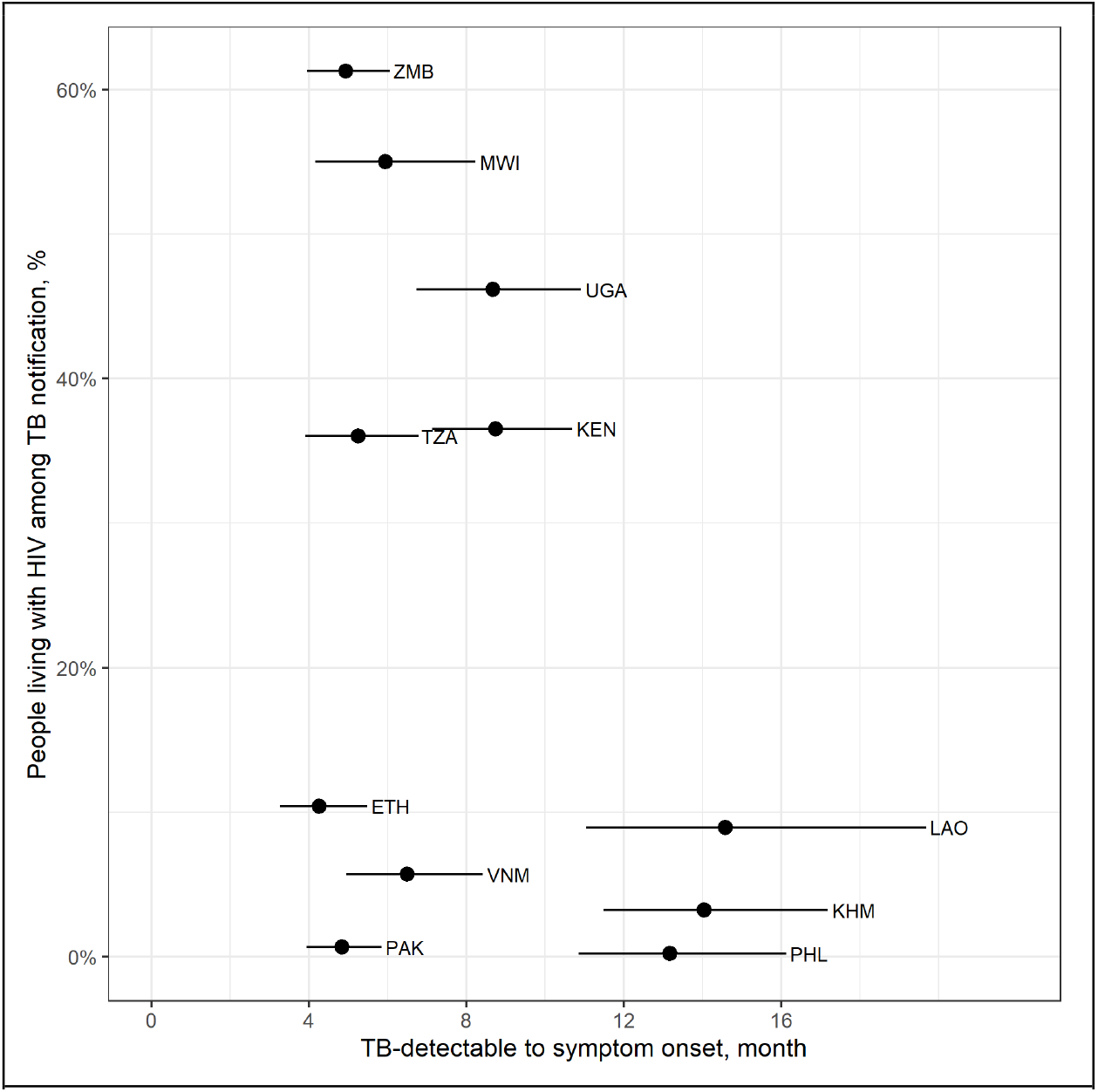
Duration from TB-detectable to symptom onset and people living with HIV among notified TB. The prevalence of people living with HIV were averaged over 2013 and 2018. Country labels: KHM=Cambodia, ETH=Ethiopia, KEN=Kenya, LAO=Lao People’s Democratic Republic, MWI=Malawi, PAK=Pakistan, PHL= Philippines, TZA=United Republic of Tanzania, UGA=Uganda, VNM=Viet Nam, ZMB=Zambia.

### Sensitivity Analyses

Across all mortality scenarios considered, differences in duration were around one month, except for Lao PDR (**Table 4**). The highest levels of assumed mortality led to up to a 7 percentage-point lower case-detection ratio than for the lowest assumed mortality. Sensitivity analysis assuming an extremely high excess TB mortality for PLHIV (**Table 3** and **Table 4-table supplement 2**) led to up to 15 percentage-point lower CDRs and correspondingly higher incidence rates. However, for both Kenya and Blantyre, the case-detection ratios were still not lower than that of HIV negative populations. Considering a broader range of symptoms resulted in shorter durations of the asymptomatic phase but longer time-spent on care-seeking given the TB prevalence survey and notification data (**Table 4-table supplement 3**).

**Table 4.**
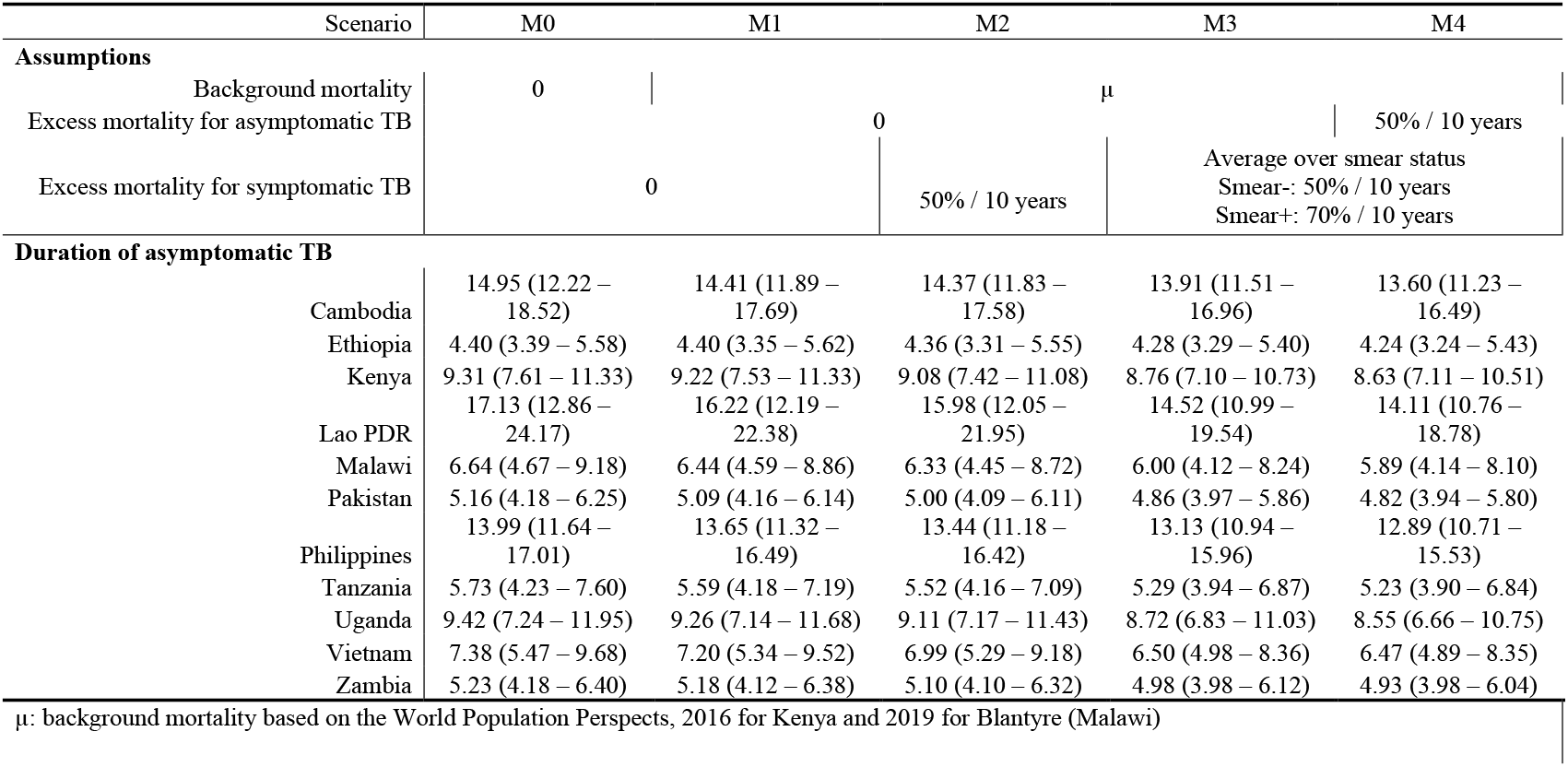
Sensitivity analysis on the asymptomatic period estimates

## Discussion

By calibrating simple models of TB disease and care-seeking progression to prevalence survey and notification data, we were able to infer the typical duration of asymptomatic bacteriologically-positive TB to be around six months. However, there is large variation between settings, with longer durations of asymptomatic disease in the included Asian settings. The asymptomatic phase typically comprised around half the total time before notification. For countries that reported care-seeking history in their prevalence surveys, we were able to estimate the average timing of initial care-seeking, finding this was approximately halfway between symptom onset and ultimate diagnosis. We found limited evidence of age-dependence in overall durations, with a hint of longer durations in older age groups, but did find substantially shorter disease durations in people living with HIV. Our analysis of TB care-seeking, diagnosis and treatment outcome cascades also showed substantial differences between settings, and meaningful losses before symptom onset and care-seeking. Taken together, these findings suggest that important opportunities exist to identify people with tuberculosis earlier in their disease course through screening and community-based active case finding interventions, potentially improving individual outcomes and reducing transmission.

TB disease prior to care-seeking, including the asymptomatic phase, is beyond the reach of passive case-finding: improvements in diagnosis and retention at the clinic will not shorten these delays nor avert deaths during this phase. Cases who are truly asymptomatic can only be found by active approaches to screening based on exposure, chest X-ray or other novel rule-in tests - based in either the community or clinics. The symptomatic period prior to care seeking would be amenable to intervention by symptom-based screening approaches and potentially health-messaging or improved access to care. Understanding the duration of these phases and their contributions to transmission and mortality is therefore key to understanding the relative potential benefits of more active approaches to finding cases [27]. Although, between 25% and 50% of the total duration (and likely a higher proportion of transmission and pre-treatment mortality) occurs once those with TB have started seeking care, when improvements in passive case-finding and reductions in pre-treatment loss-to-follow-up (which has been observed a 13% in Asia and 18% in African settings) can help [28]. While there is considerable consistency between settings in our results, the reasons for the longer durations of asymptomatic disease in Asian settings are not clear, and seem not to directly relate to HIV prevalence. These may include cultural factors around recognition of disease or willingness to report it, differences in strains, or differences in cough associated with smoking or air quality which may mask TB symptoms. It is noteworthy that these settings also have longer total durations of TB disease.

Our method also generates estimates of TB incidence: using literature estimates of TB disease mortality and self-cure rates, we inferred the proportions that fail to reach notification. Our incidence estimates are similar to those of WHO, which is not surprising since these are based on the same data. Our estimates have lower uncertainty, which may reflect under-estimation of uncertainty by our approach in not including uncertainty in TB mortality rates.

Our approach has quantified aspects of TB natural history that are potentially important but poorly characterised, relying on historical or anecdotal data. For example, mathematical models of TB transmission have from the outset often included progression in smear status [29]. In analysis of cross-sectional data, this smear conversion rate is confounded with the proportion of individuals who are smear-positive from very early in their disease. Our posterior shows high correlation between these parameters, but the gradient of initial smear-positivity with respect to conversion rate is broadly consistent between settings. Our pooled estimate should be useful for modellers wishing to parametrize joint uncertainty around these features.

There has been an increased realisation that TB disease is less dichotomous and more dynamic than conventional paradigms have allowed [8]. Our findings complement work in this area by describing the dynamics through asymptomatic disease and different levels of infectiousness while symptomatic. (We have used the term asymptomatic TB as synonymous with the term sub-clinical TB used in some discussions.) The substantial fraction of time spent asymptomatic means there is the potential for this phase to make important contributions to TB transmission; the key unknown factor being the relative infectiousness of the asymptomatic phase [9]. While some evidence suggests that the natural history of TB disease may include progression and remission [8], we have assumed a one-way progression through the asymptomatic phase, smear status, and care-seeking - we were limited here by our reliance on cross-sectional data. However our transitions between these cruder states could be considered as characterising the net average transitions in a population experiencing more complex dynamics among sub-states. Similarly, there may be heterogeneity in disease-course, which we were not able to consider.

HIV-associated TB has long been known to have a more rapid TB disease progression, but data to quantify this has been limited, and most models and estimates of TB burden assume the same case detection ratio in PLHIV as among HIV-negative people. We were able to provide the first empirical estimate of this quantity. Our sensitivity analysis considered from zero excess TB/HIV mortality through to an extremely high level, and consistently found TB case-detection ratios for PLHIV that were higher than for HIV-negative people. We were not able to stratify by ART status, but increases in ART coverage and regular healthcare visits for people on ART may have helped improve TB case detection among PLHIV. We found an asymptomatic TB disease duration shorter than five months in people living with HIV, and the delays from symptom onset to case-detection also much shorter than those for HIV-negative people.

Comparing our model estimates of the delays from TB onset to notification with empirical estimates from P:N ratios, we find that empirical estimates systematically overestimate duration. This is because the empirical estimator implicitly assumes that all prevalent TB ends in notification, whereas in the model (and reality) self-cure and death are possible outcomes. These unobserved competing events add to the rate at which individuals cease being prevalent, thus shortening the prevalent duration. The discrepancy between empirical and model estimates is largest where the CDR is lowest. Under-notification, e.g. in settings with large private sectors can further overestimate durations.

The duration of symptomatic TB is necessarily shorter than the total duration. Empirical estimates of TB duration based on TB prevalence surveys that used symptoms as an entry point to bacteriological testing may therefore be biased. Where symptoms alone are used to rule-in, the empirical P:N ratio is measuring the duration of the symptomatic phase rather than the total duration of TB disease. The range of symptoms used can also affect estimated durations, with broader definitions shortening the duration of asymptomatic phase. All the surveys we used either applied bacteriological testing to everyone, or used chest X-ray (potentially OR symptoms) to rule-in for bacteriological testing.

A limitation of our approach is that it relies on self-report for defining whether individuals have symptoms or have begun seeking care. Stigma, fear of diagnosis, or expectations of treatment may all affect and influence participants’ willingness to report symptoms, even if they are recognised; recognition of symptoms may itself be influenced by cultural and epidemiological factors [30,31]. Recall bias may play a role in limiting the accuracy of reports of care-seeking. We were limited in our ability to detect effects that may be real, such as age-dependence in durations, and to investigate smear-status while asymptomatic, due to the relatively small numbers of TB cases found in prevalence surveys. Our projections of growth or decline rely solely on trends in notification data; where notification trends reflect changes in routine case finding performance, our projections of underlying incidence and prevalence trends may be incorrect. Finally, while most of our parameters have incorporated prior uncertainty, being treated as random variables in a Bayesian framework, we treated TB mortality and self-cure as fixed, which may under-estimate uncertainty in our estimates.

A strength of our approach is that we have adopted a rigorous Bayesian framework, which includes uncertainty in inputs and outputs and could easily be generalised for use on other national or subnational data. Unlike most approaches to P:N analyses, we do not make an assumption of equilibrium. Our assumption is the slightly more general and realistic constant rate of decline, which is an additional output.

## Conclusion

Active approaches to TB screening and case-finding should be considered in high TB burden settings, where up to 4 to 12 months with infectious bacteriologically-positive disease is spent without symptoms, and a comparable time again with symptoms before care-seeking initiation.

## Supporting information

GATHER checklist

Processed data

## Data Availability

All data are available as supplementary dataset and a compressed version can be download from the project GitHub repo.

https://github.com/TimeWz667/AsymTB.git

## Acknowledgements

For sharing additional stratified TB notification data, we would like to thank the following staff at the Division of Tuberculosis, Leprosy and Lung Disease Program (DNTLD-P), Kenya: Elizabeth Onyango (head), Aiban Rono (head of M/E section), Martin Githiomi (ICT lead).

## Figures

**Figure 1-figure supplement 1:**
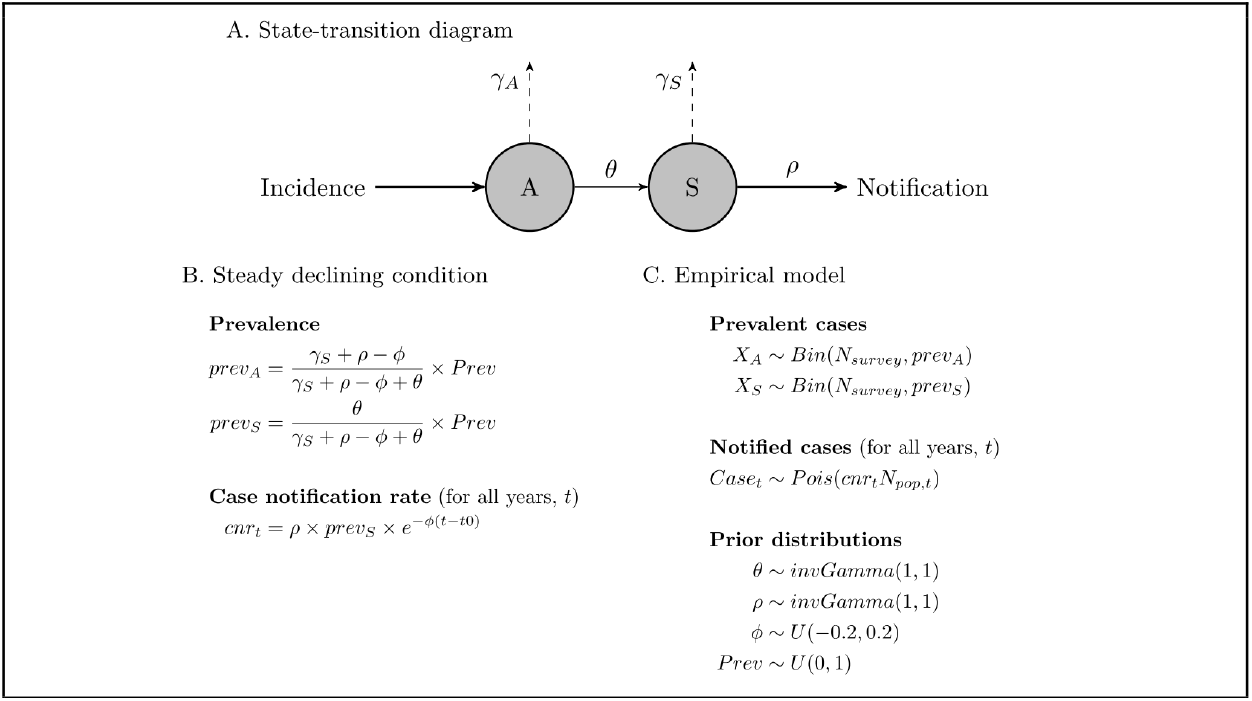
Summary of Model A. (A) State-transition diagram: *A*: asymptomatic TB; *S*: symptomatic TB. (B) steady declining condition, which has identical percentage annual decline in prevalence, incidence rate and notification rate for all states: *Prev*: overall active TB prevalence; *prev*_*A*_, *prev*_*S*_: prevalence by state; *cnr*_*t*_: case notification rate at time *t*. (C) empirical model with prior distributions: *Bin*(.): binomial distribution, *Pois*(.): Poisson distribution, *invGamma*(.): inverse-gamma distribution, *U*(.): uniform distribution. See Figure 1-table supplement 1 for the notations of parameters and data

**Figure 1-figure supplement 2:**
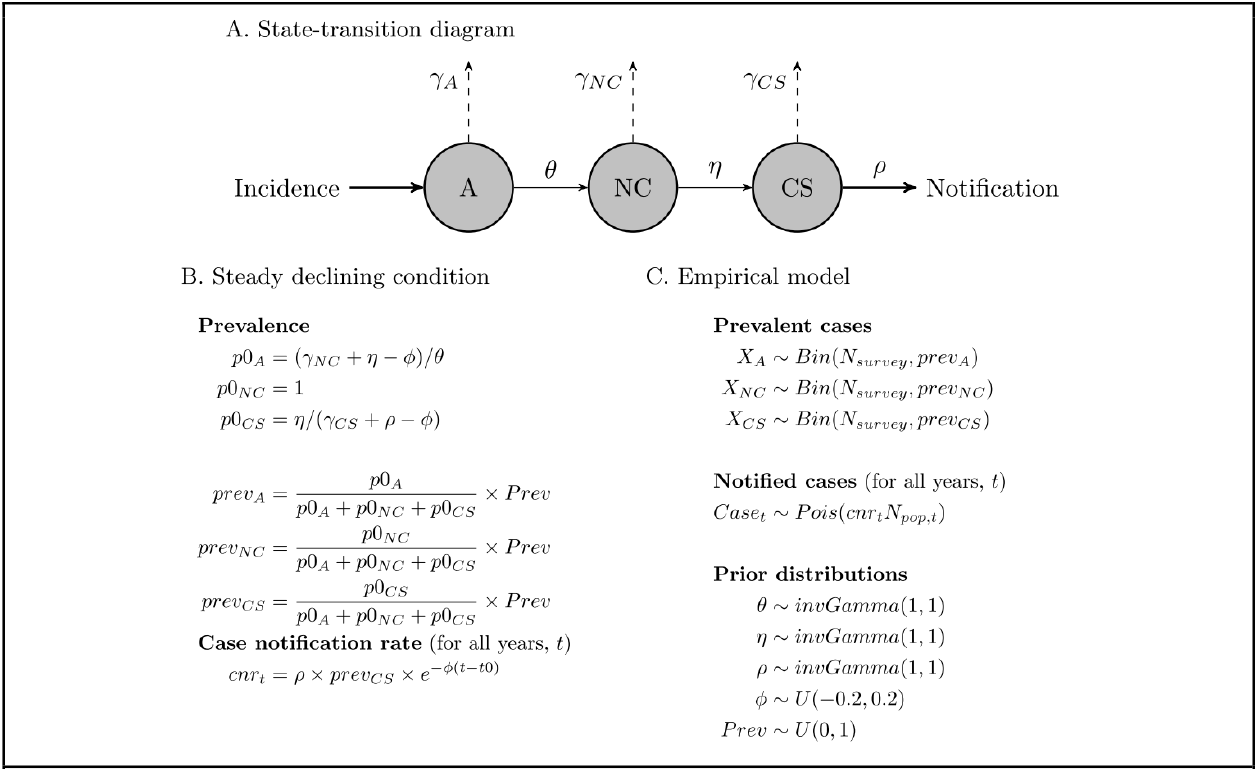
Summary of Model B. (A) State-transition diagram: *A*: asymptomatic TB; *NC*: symptomatic TB without care-seeking attempts; *CS*: symptomatic TB started care-seeking. (B) steady declining condition, which has identical percentage annual decline in prevalence, incidence rate and notification rate for all states: *Prev*: overall active TB prevalence; *prev*_*A*_, *prev*_*NC*_, *prev*_*CS*_: prevalence by state; *cnr*_*t*_: case notification rate at time *t*. (C) empirical model with prior distributions: *Bin*(.): binomial distribution; *Pois*(.): Poisson distribution; *invGamma*(.): inverse-gamma distribution; *U*(.): uniform distribution. See Figure 1-table supplement 1 for the notations of parameters and data

**Figure 1-figure supplement 3:**
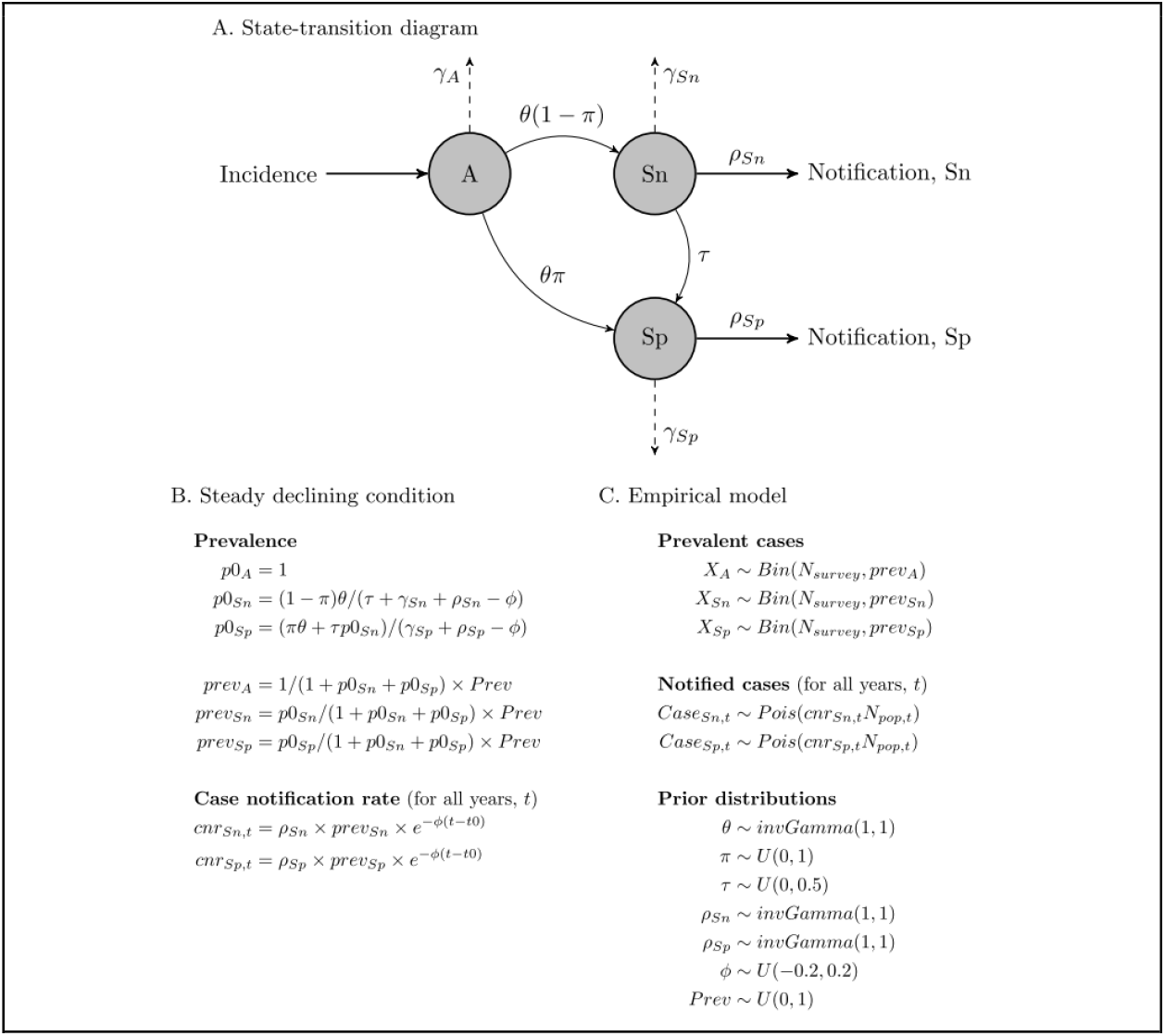
Summary of Model C. (A) State-transition diagram: *A*: asymptomatic TB; *Sn*: symptomatic TB, smear-negative; *Sp*: symptomatic TB, smear-positive. (B) steady declining condition, which has identical percentage annual decline in prevalence, incidence rate and notification rate for all states: *Prev*: overall active TB prevalence; *prev*_*A*_, *prev*_*Sn*_, *prev*_*Sp*_: prevalence by state; *cnr*_*Sn,t*_, *cnr*_*Sp,t*_: case notification rate by state at time *t*. (B) empirical model with prior distributions: *Bin*(.): binomial distribution; *Pois*(.): Poisson distribution; *invGamma*(.): inverse-gamma distribution; *U*(.): uniform distribution. See Figure 1-table supplement 1 for the notations of parameters and data

**Figure 1-table supplement 1:** Parameter notations and descriptions.

| Notation | Description | Prior distribution / Source |
| --- | --- | --- |
| Data |  |  |
| $X_s$ | Prevalent TB in state $s$ | TB prevalence surveys |
| $N_{survey}$ | Eligible subjects in TB prevalence surveys | TB prevalence surveys |
| $t_0$ | Year of TB prevalence surveys | TB prevalence surveys |
| $Case_t$ | Notified cases | WHO TB notification |
| $N_{pop,t}$ | Population size at time $t$ | WPP data |
| $\mu_s$ | Untreated death rate of state $s$ | WPP data |
| Parameters |  |  |
| $\theta$ | Symptom onset rate | $invGamma(1, 1)$ |
| $\pi$ | Proportion of smear-positive at symptom onset | $U(0, 1)$ |
| $\tau$ | Conversion rate, from smear-negative to -positive | $U(0, 0.5)$ |
| $\eta$ | Presence rate of care-seeking intentions | $invGamma(1, 1)$ |
| $\rho$ | Care-seeking rate | $invGamma(1, 1)$ |
| $\nu$ | Self-cure rate | $U(0.1, 0.3)$ |
| $\gamma_s$ | Dropout rate before case-detection | $\nu + \gamma_s$ |
| $\phi$ | Annual decline rate for incidence | $U(-0.2, 0.2)$ |

WPP: World Population Prospects 2019. $invGamma(.)$ : inverse-gamma distribution, $U(.)$ : uniform distribution

**Figure 2-figure supplement 1.**
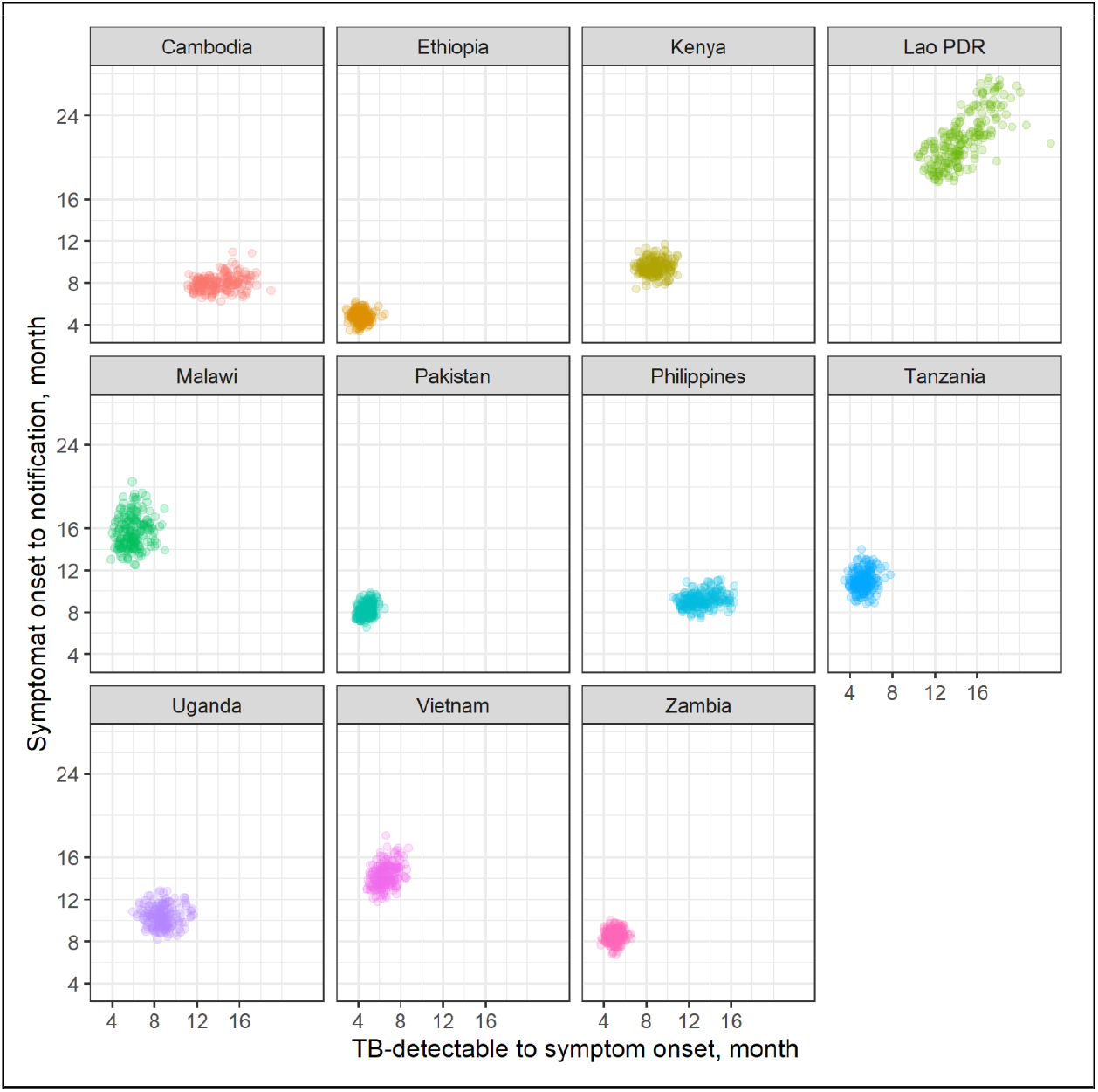
Duration from TB-detectable to symptom onset and from symptom onset to case-detection by country. Each point represents a sample from the posterior parameters.

**Figure 4-figure supplement 1.**
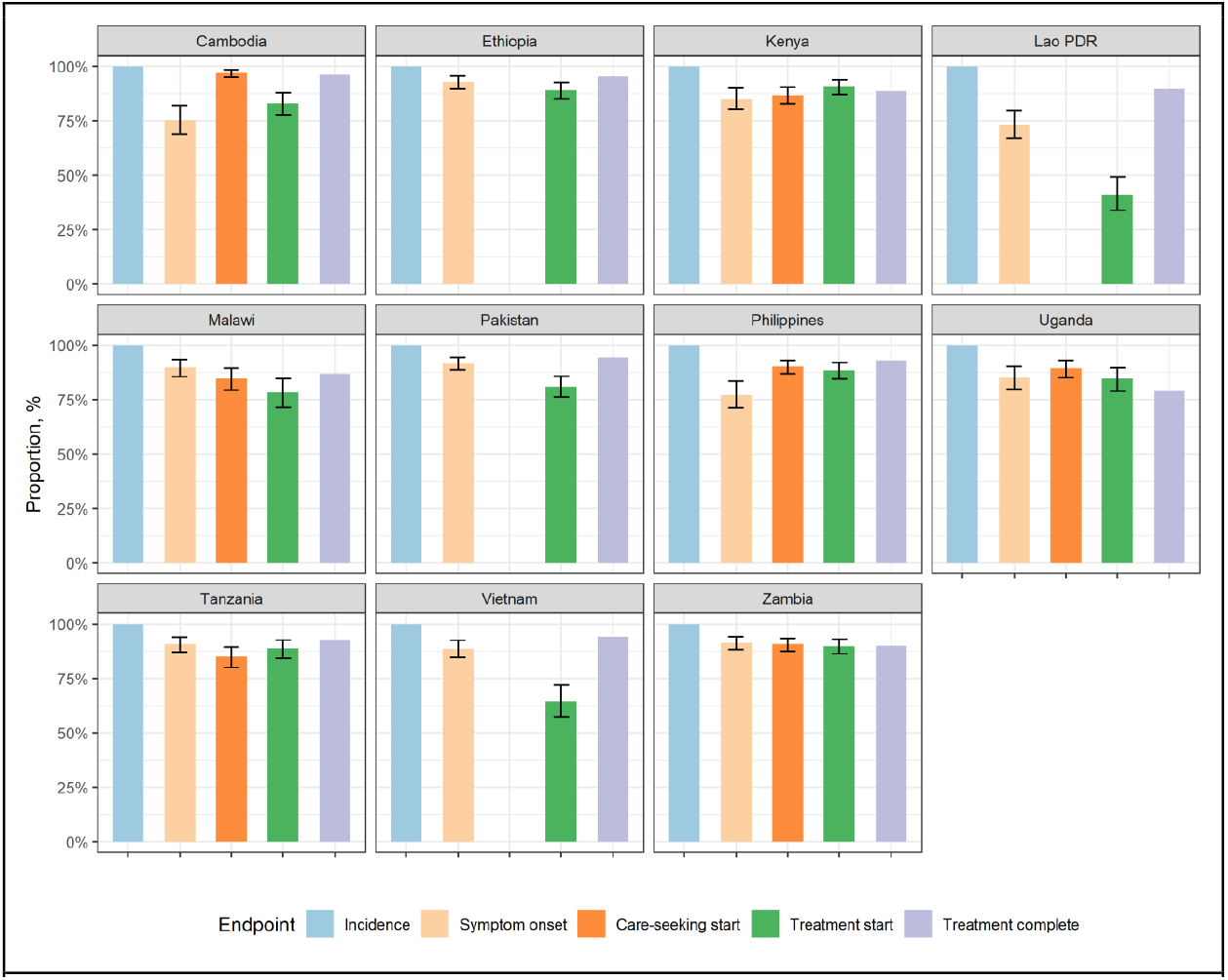
Healthcare gaps. The values are the fraction of those beginning the previous stage who reach the labelled stage. The second bars may be missing if the TB prevalence survey did not report along with the results of care-seeking behaviour. The treatment complete here were sourced from the WHO TB outcome data, including treatment completion and treatment success.

**Figure 4-figure supplement 2.**
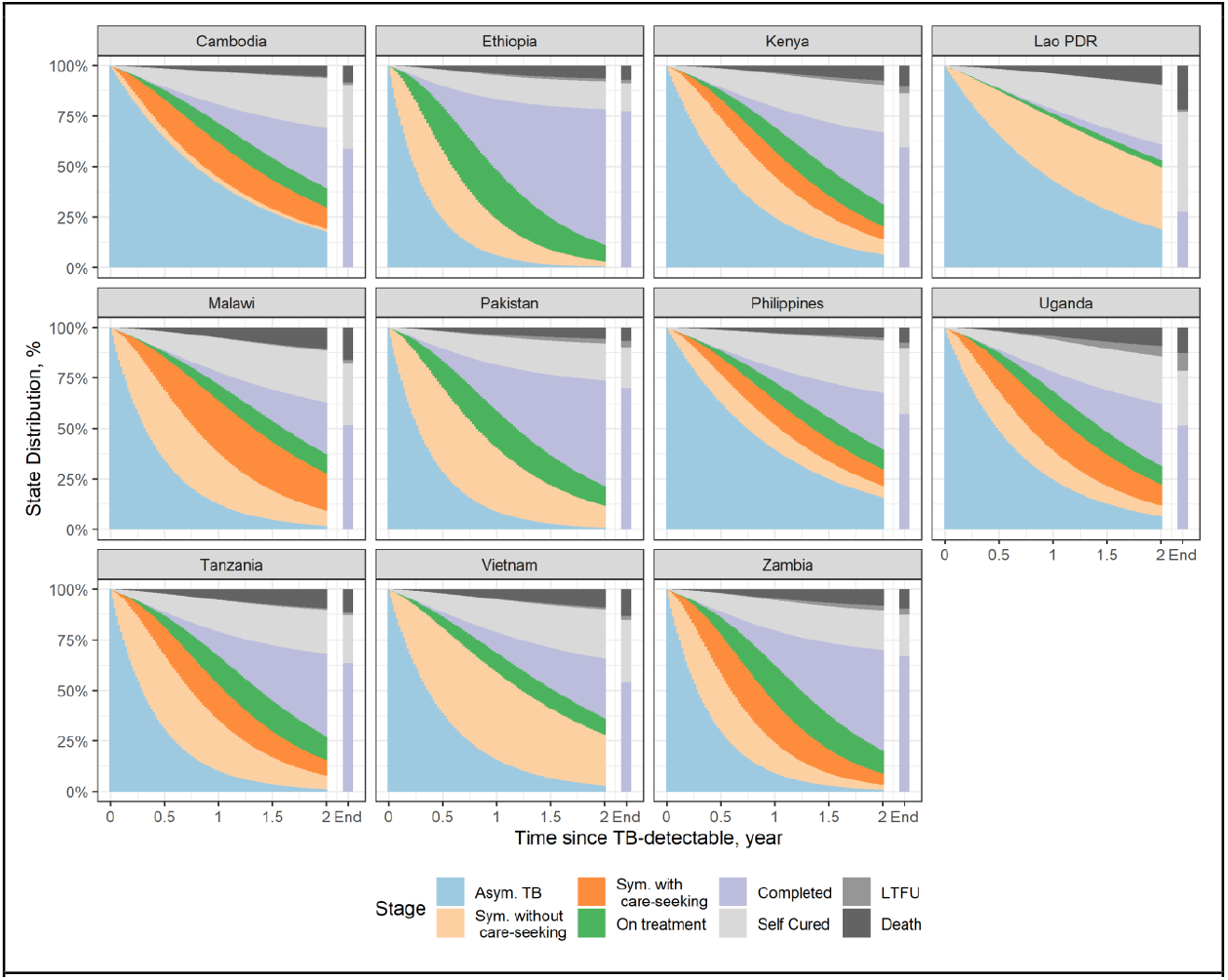
Stage distribution by time since TB detectable. The size of each area represents the time-spent in each stage. The treatment complete here were sourced from the WHO TB outcome data, including treatment completion and treatment success; LTFU: loss to follow-up.

**Figure 5-figure supplement 1.**
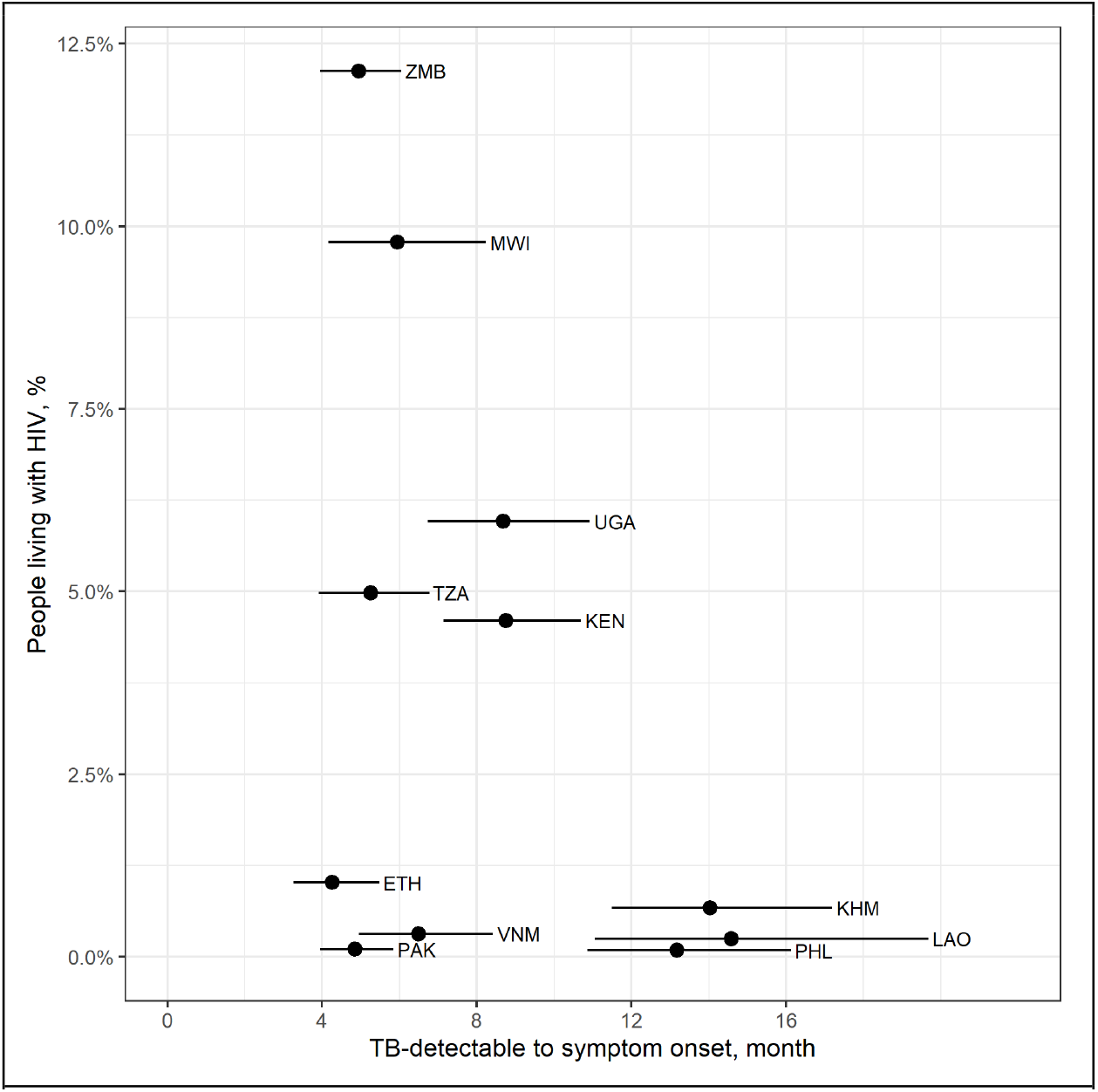
Duration from TB-detectable to symptom onset and people living with HIV, aged 15 and above. The prevalence of people living with HIV were averaged over 2013 and 2018. Country labels: KHM=Cambodia, ETH=Ethiopia, KEN=Kenya, LAO=Lao People’s Democratic Republic, MWI=Malawi, PAK=Pakistan, PHL= Philippines, TZA=United Republic of Tanzania, UGA=Uganda, VNM=Viet Nam, ZMB=Zambia.

**Figure 5-figure supplement 2.**
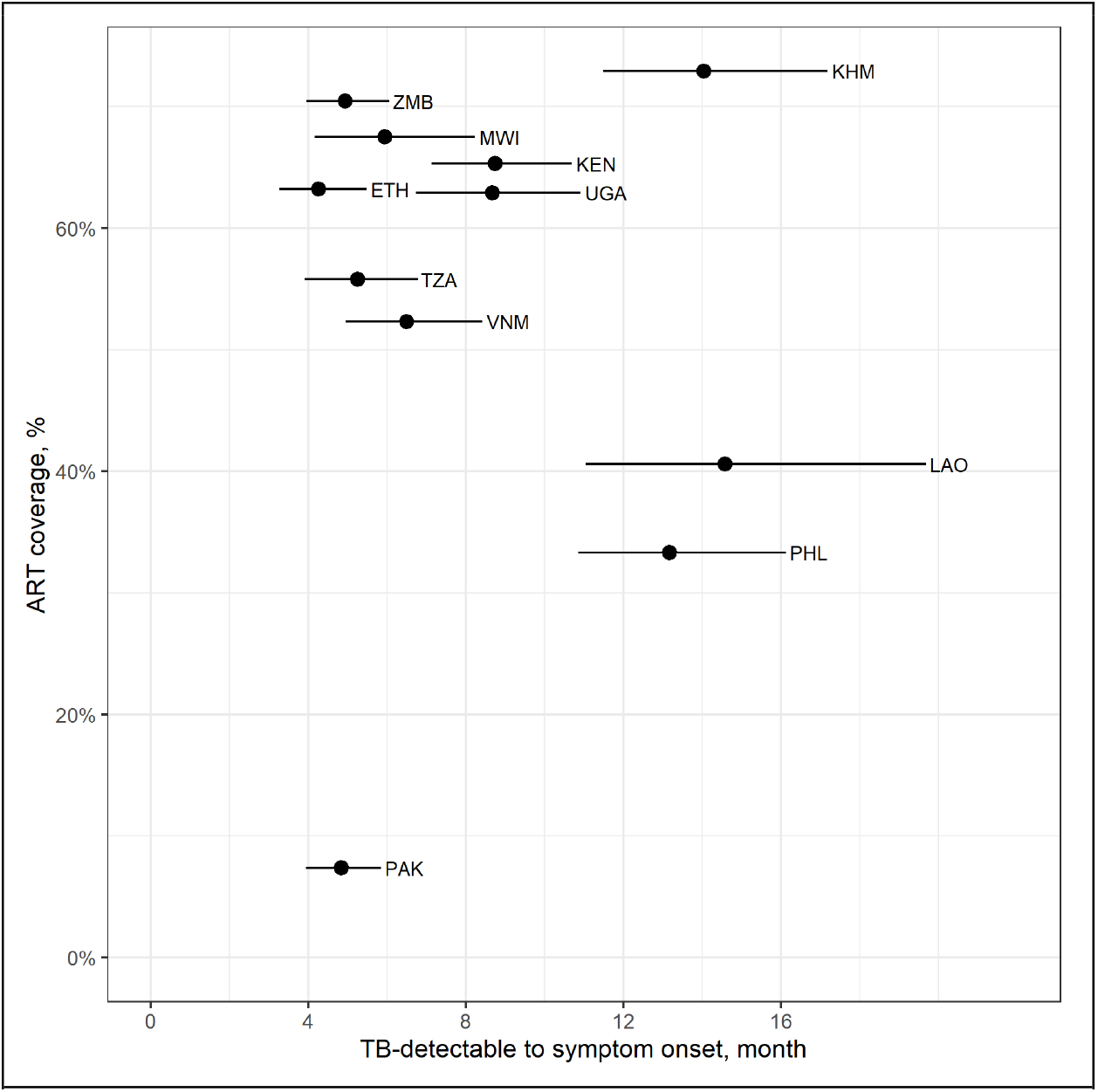
Duration from TB-detectable to symptom onset and ART coverage, aged 15 and above. The ART coverage were averaged over 2013 and 2018. Country labels: KHM=Cambodia, ETH=Ethiopia, KEN=Kenya, LAO=Lao People’s Democratic Republic, MWI=Malawi, PAK=Pakistan, PHL= Philippines, TZA=United Republic of Tanzania, UGA=Uganda, VNM=Viet Nam, ZMB=Zambia.

## Tables

**Table 1. table supplement 1.** Extracted information from the national TB prevalence surveys

| Country | Survey year | Reported prevalence estimates,<br>per 100 000 with 95% confidence<br>interval | Cases with TB suggestive<br>symptoms | Symptoms | Raw counts in the analysis<br>(Asymptomatic + Symptomatic) / subjects | Care-seeking behaviour<br>reported |
| --- | --- | --- | --- | --- | --- | --- |
| Cambodia | 2011 | Overall: 831 (707-977)<br>Female: 609 (486-763)<br>Male: 1,097 (895-1343) | 23% of Smr-<br>44% of Smr+ | Cough ≥ 2 weeks and<br>haemoptysis | Total: (221 + 93) / 37,417<br>Female: (89 + 37) / 20,410<br>Male: (132 + 56) / 17,007 | Survey participants with<br>screening symptoms<br>Female: 92%<br>Male: 84% |
| Ethiopia | 2010-2011 | Overall: 277 (208-347)<br>Female: 246 (176-315)<br>Male: 304 (219-388) | 48% of Smr-<br>57% of Smr+ | Cough ≥ 2 weeks | Total: (53 + 57) / 46,697<br>Female: (26 + 28) / 24,878<br>Male: (27 + 29) / 21,819 | |
| Kenya | 2016 | Overall: 558 (455-662)<br>Female: 359 (258-460)<br>Male: 809 (656-962) | Exact counts | Cough ≥ 2 weeks | Total: (161 + 140) / 62,988<br>Female: (62 + 52) / 36,974<br>Male: (99 + 88) / 26,014 | Survey TB cases with<br>symptoms<br>Female: 21 (40%)<br>Male: 32 (36%) |
| Lao PDR | 2010-2011 | Overall: 595 (457-733)<br>Female: 366 (254-477)<br>Male: 855 (646-1064) | 36% of Smr-<br>66% of Smr+ | Cough ≥ 2 weeks or<br>haemoptysis | Total: (119 + 118) / 39,212<br>Female: (41 + 39) / 21,474<br>Male: (78 + 79) / 17,738 | |
| Malawi | 2013 | Total: 452 (312-593)<br>Female: 374 (246-501)<br>Male: 546 (335-757) | 73% of Smr-<br>66% of Smr+ | Cough ≥ 2 weeks | Total: (40 + 92) / 31,579<br>Female: (21 + 48) / 18,490<br>Male: (19 + 44) / 13,089 | Survey TB cases with<br>symptoms<br>Smr-: 51%<br>Smr+: 61% |
| Pakistan | 2010-2011 | Total: 398 (333-463)<br>Female: 320 (253-388)<br>Male: 484 (392-577) | 61% of all BC | Cough ≥ 2 weeks | Total: (132 + 208) / 104,631<br>Female: (62 + 98) / 60,382<br>Male: (70 + 110) / 44,249 | |
| Philippines | 2016 | Overall: 1,159 (1,016-1,301)<br>Female: 627 (516-739)<br>Male: 1,713 (1,482-1,943) | 21% of Smr-<br>51% of Smr+ | Cough ≥ 2 weeks and<br>haemoptysis | Total: (316 + 150) / 46,689<br>Female: (99 + 45) / 25,796<br>Male: (217 + 105) / 20,893 | Survey TB cases with<br>symptoms<br>Total: 71% |
| Uganda | 2014-2015 | Total: 401 (292-509)<br>Female: 178 (109-248)<br>Male: 734 (554-914) | 46% of Smr-<br>52% of Smr+ | Cough ≥ 2 weeks | Total: (83 + 77) / 40,539<br>Female: (21 + 19) / 23,393<br>Male: (62 + 58) / 17,146 | Survey participants with<br>screening symptoms<br>Female: 67%<br>Male: 54% |
| Tanzania | 2012 | Total: 293 (228-358)<br>Female: 204 (137-272)<br>Male: 410 (315-505) | 63% of all BC | Cough ≥ 2 weeks,<br>haemoptysis, fever ≥ 2<br>weeks, weight loss and<br>excessive night sweats | Total: (56 + 98) / 50,436<br>Female: (23 + 41) / 29,701<br>Male: (33 + 57) / 20,735 | Survey participants with<br>screening symptoms<br>Female: 39.1%<br>Male: 37.7% |
| Vietnam | 2017-2018 | Total: 322 (260-399)<br>Female: 133 (89-198)<br>Male: 522 (420-648) | 67% of all BC | Cough ≥ 2 weeks | Total: (73 + 148) / 61,763<br>Female: (15 + 31) / 34,613<br>Male: (58 + 117) / 27,150 | |
| Zambia | 2013-2014 | Total: 638 (502-774)<br>Female: 487 (353-621)<br>Male: 833 (641-1,024) | 61% of all BC | Cough ≥ 2 weeks | Total: (105 + 160) / 46,099<br>Female: (45 + 68) / 26,642<br>Male: (60 + 92) / 19,457 | Survey TB cases with<br>symptoms<br>Total: 50% |

**Table 4-table supplement 1 Sensitivity analysis:**
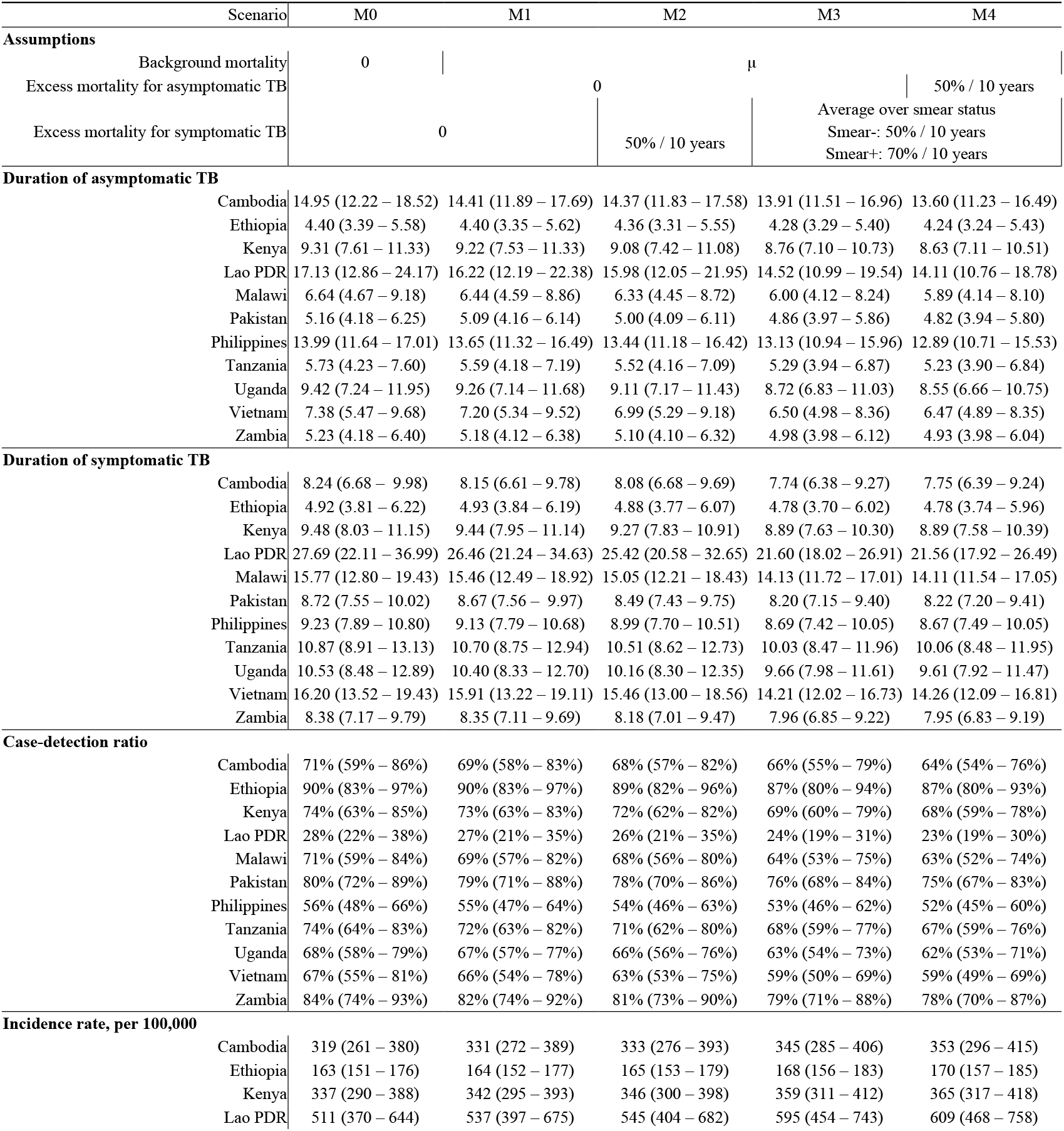
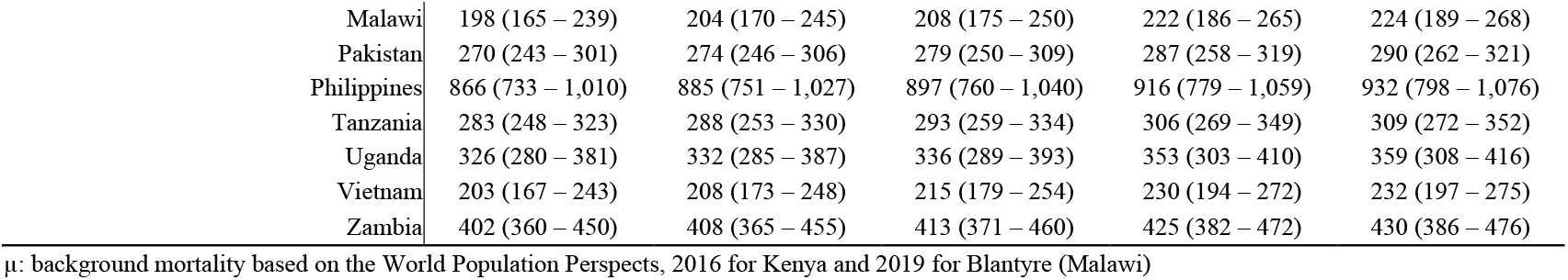
assumptions on untreated TB mortality rates

**Table 4-table supplement 2 Sensitivity analysis:** assumptions on untreated TB mortality rates for people living with HIV

|  | Blantyre |  |  |  | Kenya |  |  |  |
| --- | --- | --- | --- | --- | --- | --- | --- | --- |
| HIV status | Non-HIV | PLHIV |  |  | Non-HIV | PLHIV |  |  |
| Assumptions |  |  |  |  |  |  |  |  |
| Background mortality | μ | μ + HIV-related mortality |  |  | μ | μ + HIV-related mortality |  |  |
| excess mortality for symptomatic TB | Smear-: 50% / 10 years<br>Smear+: 70% / 10 years | 0 | Smear-: 50% / 10 years<br>Smear+: 70% / 10 years | 5% per month | Smear-: 50% / 10 years<br>Smear+: 70% / 10 years | 0 | Smear-: 50% / 10 years<br>Smear+: 70% / 10 years | 5% per month |
| All TB regardless of symptomatic or not |  |  |  |  |  |  |  |  |
| Time to symptom onset, month | 8.79 (5.14 – 13.18) | 1.27 (0.27 – 2.91) | 1.32 (0.31 – 3.04) | 1.15 (0.25 – 2.64) | 10.35 (8.41 – 12.84) | 3.83 (2.42 – 5.50) | 3.81 (2.34 – 5.51) | 3.22 (1.99 – 4.75) |
| Time to notification, month | 16.27 (11.59 – 21.53) | 3.76 (1.85 – 6.16) | 3.76 (1.81 – 6.07) | 3.26 (1.76 – 5.03) | 20.90 (18.01 – 24.46) | 8.03 (5.92 – 10.36) | 7.79 (5.78 – 9.99) | 6.49 (4.94 – 8.11) |
| Case-detection ratio | 76% (64% – 88%) | 95% (90% – 98%) | 94% (88% – 98%) | 84% (74% – 93%) | 69% (59% – 81%) | 86% (78% – 92%) | 84% (77% – 91%) | 71% (63% – 80%) |
| Incidence rate, per 100,000 | 158 (132 – 192) | 1,718 (1,608 – 1,845) | 1,742 (1,618 – 1,879) | 1,949 (1,745 – 2,217) | 263 (221 – 306) | 1,987 (1,846 – 2,166) | 2,025 (1,873 – 2,205) | 2,403 (2,142 – 2,715) |
| All symptomatic TB |  |  |  |  |  |  |  |  |
| Time to notification, month | 7.47 (4.53 – 11.05) | 2.49 (1.03 – 4.40) | 2.44 (1.00 – 4.25) | 2.11 (0.95 – 3.49) | 10.56 (8.86 – 12.48) | 4.20 (2.65 – 6.05) | 3.99 (2.52 – 5.56) | 3.27 (2.16 – 4.40) |
| Case-detection ratio | 88% (79% – 95%) | 97% (93% – 99%) | 95% (90% – 98%) | 85% (75% – 94%) | 83% (75% – 91%) | 92% (87% – 96%) | 90% (85% – 95%) | 76% (67% – 84%) |
| Incidence rate, per 100,000 | 136 (121 – 155) | 1,690 (1,594 – 1,802) | 1,712 (1,604 – 1,831) | 1,921 (1,721 – 2,177) | 218 (198 – 240) | 1,845 (1,765 – 1,958) | 1,882 (1,791 – 2,003) | 2,260 (2,025 – 2,542) |

μ: background mortality based on the World Population Perspects, 2016 for Kenya and 2019 for Blantyre (Malawi)
HIV-related mortality: AIDS-related deaths / PLHIV for aged 15 and above from the UNAIDS data
5% per month (0.61 per year): an extreme case for gold miner in pre-ART era used by []

**Table 4-table supplement 3 Sensitivity analysis:** definition of symptomatic TB

| Symptomatic | Blantyre |  | Kenya |  |  |
| --- | --- | --- | --- | --- | --- |
|  | Cough >= 2 weeks | Definition 2 | Cough >= 2 weeks | Definition 2 | Definition 3 |
| **All TB regardless of symptomatic or not** |  |  |  |  |  |
| Time to symptom onset, month | 4.45 (2.70 – 6.65) | 2.96 (1.56 – 4.73) | 8.80 (7.17 – 10.68) | 6.00 (4.71 – 7.53) | 1.43 (0.93 – 2.09) |
| Time to notification, month | 8.87 (6.42 – 11.67) | 8.55 (6.20 – 11.16) | 17.74 (15.37 – 20.41) | 17.32 (15.06 – 20.04) | 16.25 (13.96 – 18.91) |
| Case-detection ratio | 85% (77% – 93%) | 85% (77% – 92%) | 69% (59% – 79%) | 68% (59% – 78%) | 68% (58% – 77%) |
| Incidence rate, per 100,000 | 382 (346 – 427) | 383 (347 – 427) | 358 (311 – 414) | 363 (317 – 417) | 365 (321 – 421) |
| **All symptomatic TB** |  |  |  |  |  |
| Time to notification, month | 4.40 (2.50 – 6.39) | 5.60 (3.71 – 7.78) | 8.91 (7.61 – 10.39) | 11.30 (9.80 – 13.09) | 14.81 (12.69 – 17.29) |
| Case-detection ratio | 91% (85% – 96%) | 89% (81% – 94%) | 80% (73% – 87%) | 75% (68% – 83%) | 69% (60% – 78%) |
| Incidence rate, per 100,000 | 357 (334 – 387) | 367 (340 – 401) | 308 (284 – 337) | 328 (298 – 364) | 357 (317 – 408) |

Definition 2: cough >= 14 days, hemoptysis, fever, weight loss, night sweats
Definition 3: cough >= 14 days, hemoptysis, fever, breathe shortness, hemoptysis, chest pains, night sweats, weight loss

