## Supplementary material for "Estimated durations of asymptomatic, symptomatic, and care-seeking phases of tuberculosis disease": GATHER checklist

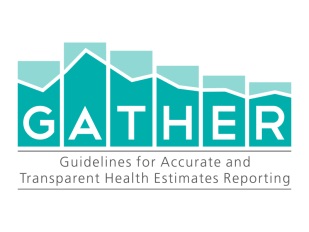
**Checklist of information that should be included in new reports of global health estimates**

| Item # | Checklist item | Reported on page # |
| --- | --- | --- |
| Objectives and funding | | |
| 1 | Define the indicator(s), populations (including age, sex, and geographic entities), and time period(s) for which estimates were made. | I5, M1-3 |
| 2 | List the funding sources for the work. | Financial statement |
| Data Inputs | | |
| *For all data inputs from multiple sources that are synthesized as part of the study:* | | |
| 3 | Describe how the data were identified and how the data were accessed. | M2-3 |
| 4 | Specify the inclusion and exclusion criteria. Identify all ad-hoc exclusions. | M1 |
| 5 | Provide information on all included data sources and their main characteristics. For each data source used, report reference information or contact name/institution, population represented, data collection method, year(s) of data collection, sex and age range, diagnostic criteria or measurement method, and sample size, as relevant. | M5-8, R1 |
| 6 | Identify and describe any categories of input data that have potentially important biases (e.g., based on characteristics listed in item 5). | M10-12, M14 |
| *For data inputs that contribute to the analysis but were not synthesized as part of the study:* | | |
| 7 | Describe and give sources for any other data inputs. | M4-8 |
| *For all data inputs:* | | |
| 8 | Provide all data inputs in a file format from which data can be efficiently extracted (e.g., a spreadsheet rather than a PDF), including all relevant meta-data listed in item 5. For any data inputs that cannot be shared because of ethical or legal reasons, such as third-party ownership, provide a contact name or the name of the institution that retains the right to the data. | Source Data |
| Data analysis | | |
| 9 | Provide a conceptual overview of the data analysis method. A diagram may be helpful. | M9-11  Appendix 1 |
| 10 | Provide a detailed description of all steps of the analysis, including mathematical formulae. This description should cover, as relevant, data cleaning, data pre-processing, data adjustments and weighting of data sources, and mathematical or statistical model(s). | M9-12 |
| 11 | Describe how candidate models were evaluated and how the final model(s) were selected. | M9, M12 |
| 12 | Provide the results of an evaluation of model performance, if done, as well as the results of any relevant sensitivity analysis. | Table 4 |
| 13 | Describe methods for calculating uncertainty of the estimates. State which sources of uncertainty were, and were not, accounted for in the uncertainty analysis. | M12-14 |
| 14 | State how analytic or statistical source code used to generate estimates can be accessed. | M12, G |
| Results and Discussion | | |
| 15 | Provide published estimates in a file format from which data can be efficiently extracted. | G |
| 16 | Report a quantitative measure of the uncertainty of the estimates (e.g. uncertainty intervals). | R2, R4-6 |
| 17 | Interpret results in light of existing evidence. If updating a previous set of estimates, describe the reasons for changes in estimates. | R2, D3-4 |
| 18 | Discuss limitations of the estimates. Include a discussion of any modelling assumptions or data limitations that affect interpretation of the estimates. | D5-6, D9-10 |

*This checklist should be used in conjunction with the GATHER statement and Explanation and Elaboration document, found on gather-statement.org*

I: Introduction, M: methods, R: results, D: discussion, Fig: figure, Tab: table

G: GitHub repo of TimeWz667/AsymTB

The numbers specified paragraphs. E.g. I3 is the 3^rd^ paragraph of introduction section.
